# Plasma proteomics of *APOE* genotype: age-specific analyses in UK population-based cohorts

**DOI:** 10.64898/2026.04.16.26351010

**Authors:** Amy Packer, Tahsina Khatun, James W Groves, Tony Wyss-Coray, Jonathan M Schott, Petroula Proitsi, Emma L Anderson, Dylan M Williams

## Abstract

**Background:** The apolipoprotein E (*APOE*) locus is the strongest genetic risk factor for late-onset Alzheimer’s disease (AD). Variation in *APOE* isoforms is known to have diverse pleiotropic effects on circulating lipids and other metabolites, but effects on the circulating proteome across the life course are not well characterised. We investigated the specific effects of *APOE* ε4 and *APOE* ε2 carriage on the circulating proteome in middle-age and later life.

**Methods:** In primary modelling, we analysed associations of *APOE* ε4 and ε2 carriage (reference ε3/ε3) with circulating proteins in UK Biobank participants (*N* = 42,642; age = 39.1 to 70.9 years). Using multivariable linear regression, we conducted ancestry-specific analyses of 2,922 assayed plasma proteins across individuals of European (EUR), African (AFR), and South Asian (SAS) ancestry. To identify age-dependent effects, stratified analyses were performed with the sample split into age groups. We then performed replication analyses of *APOE*-associated proteins in age-matched groups, using data from two independent UK-based cohorts.

**Results:** We identified 351 proteins associated with ε2 carriage and 480 with ε4 carriage among individuals of European ancestry (*n* = 40,092); 130 of these were associated with both ε2 and ε4 carriage (with either consistent or inverse association directions). These included established biomarkers of neurodegeneration (GFAP and NEFL) and other proteins implicated by AD genetic risk loci (e.g., TREM2, CTSB, IDUA, SORT1, GRN). Many of these proteins are linked to other neurodegenerative diseases besides AD. In multiple age groups, ε4 carriage was strongly associated with consistent differences in circulating APOE, MENT, and PLA2G7 levels across ancestries and cohorts.

**Conclusion:** *APOE* ε4 and ε2 exert broad, often age-dependent effects on the plasma proteome, detectable decades before typical ages of AD diagnoses, highlighting a potential early window for monitoring and intervention.

## Background

The apolipoprotein E (*APOE*) locus is the strongest genetic risk factor for late-onset dementia, including Alzheimer’s disease (AD), the most common cause of dementia (Neuropathology Group. Medical Research Council Cognitive Function and Aging Study, 2001). Three common *APOE* alleles - ε2, ε3, and ε4 - encode distinct APOE isoforms, with differential effects on risk of late-onset AD. Relative to the most common ε3 allele, ε4 significantly increases risk, whereas risk is much lower with ε2 carriage (Raulin et al., 2022). Crucially, mitigating the harmful effects of the ε3 and ε4 isoforms could prevent most AD cases and a significant proportion of all-cause dementia (Williams et al., 2026). Despite this, knowledge of when and how *APOE* variation confers AD risk remains limited, and improved characterisation of APOE’s molecular effects may help to elucidate pathways linking the protein to AD pathophysiology.

Existing molecular epidemiology to characterise APOE’s effects has largely focused on the impact of *APOE* ε4 on lipid metabolism, reflecting the primary role of APOE as a lipid transporter (Yang et al., 2023). Less attention has been given to the broader proteomic consequences of *APOE* variation. Recent advances in high-throughput proteomic profiling now enable systematic interrogation of thousands of circulating proteins for a given sample matrix (Cui et al., 2022), offering opportunities to better understand pathophysiological mechanisms, identify potential drug targets, and highlight analytes predictive of AD risk. However, understanding how *APOE* variation influences the proteome across the life course remains limited, with most research focused on older individuals and AD cases, despite neuropathological and physiological changes beginning decades before clinical symptoms (Frisoni et al., 2010; Jack Jr. et al., 2024).

In this study, we characterise the effects of *APOE* ε4 and ε2 carriage on the plasma proteome, relative to ε3 homozygosity, across middle- and older-aged participants of large UK population-based cohorts.

## Methods

Figure 1 provides an overview of the study design.

**Figure 1.**
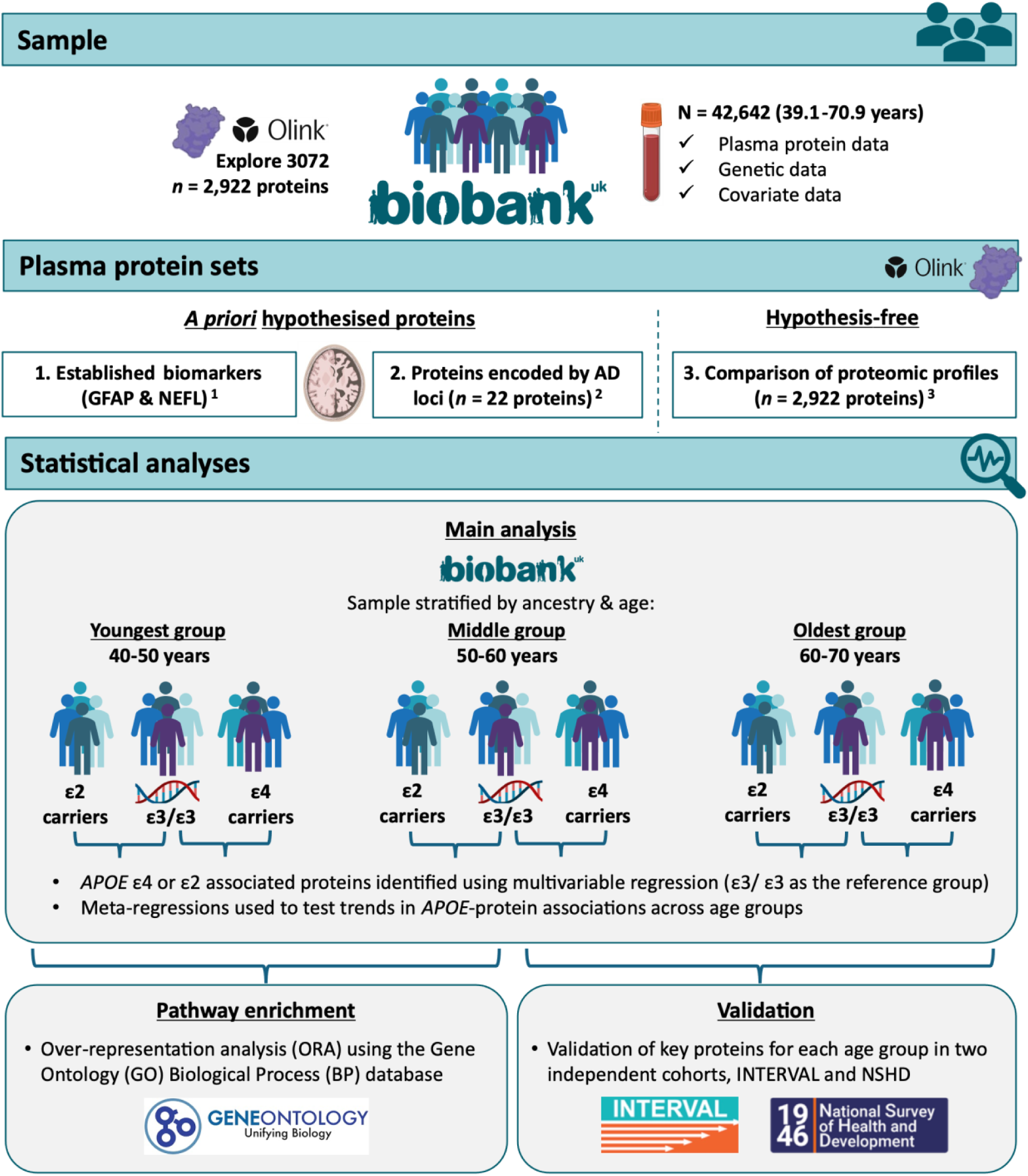
Study overview. ***Note.*** *APOE* = Apolipoprotein E; AD = Alzheimer’s disease; GFAP = glial fibrillary acidic protein; and NEFL (clinically, NfL) = neurofilament light. This figure provides an overview of the study design and data used ^1^ Biomarkers of neurodegeneration highlighted as having potential for clinical use for Alzheimer’s disease and related disorders (Hansson et al., 2023; Zetterberg & Schott, 2022) ^2^ Identified by the most recent and largest genome-wide association study at the time of analysis (Bellenguez et al., 2022) ^3^ After UKB-PPP and study-specific quality control (Sun et al., 2023).

### Sample

UK Biobank (UKB) is a prospective cohort of ∼502,000 adults aged 39-73 years at recruitment with deep phenotypic and genetic data collected (Bycroft et al., 2018). Full details of the UKB design, participants, quality control and its strengths and limitations have been described previously (Bycroft et al., 2018; Fry et al., 2017; Schoeler et al., 2023). Ethical approval was received from the North-West Multi-centre Research Ethics Committee (REC reference numbers 11/NW/0382 and 16/NW/0274).

Proteomic profiling was conducted on plasma samples from 54,219 participants through the UK Biobank Pharma Proteomics Project (UKB-PPP) (Sun et al., 2023). After restricting to individuals with quality-controlled proteomic, whole-exome sequencing, and genotype data and excluding related individuals (third-degree relatives or closer) (Manichaikul et al., 2010; *Sam Choi / GreedyRelated · GitLab*, 2020), 42,642 participants across ancestries were available. European ancestry was defined as participants who self-identified as ‘White British’ with similar genetic ancestry based on a principal components analysis of the genotypes (UKB data field ID 22006). The remaining ancestral groups were defined as described previously (Meng et al., 2024). Primary analyses focused on individuals of European ancestry (*n* = 40,092) owing to a larger sample size, with additional ancestry-specific analyses performed for African (AFR; *n* = 1,335) and South Asian (SAS; *n* = 971) individuals. We lacked sufficient power to analyse individuals of East Asian ancestry (EAS; *n* = 244). Further information, detailed inclusion criteria, and quality control procedures are described in Supplementary Methods 1.

### *APOE* coding

*APOE* alleles (ε2, ε3, ε4) were derived from single-nucleotide polymorphisms rs7412 and rs429358 (Wenham et al., 1991) using whole-exome sequencing data (Van Hout et al., 2020). Separate binary variables were created for ε2 and ε4 carriage, with non-carriers defined as ε3 homozygotes; ε2/ε4 individuals were excluded. We used ε3 homozygotes as the reference group in analyses because ε3/ε3 is the most common genotype (Auton et al., 2015).

### Proteomic measurement, processing and quality control

Plasma proteins were measured using the Olink Explore 3072 proximity extension assay, as described previously (Sun et al., 2023). Data were provided as normalized protein expression (NPX) values, Olink’s arbitrary unit of relative protein quantification on a log_2_ scale. Higher values represent higher protein concentrations. Each protein is labelled by Olink with an identifier, which typically corresponds to the gene encoding that protein (linked to UniProt IDs), rather than the name of the protein itself. Following UKB-PPP and study-specific quality control (see Supplementary Method 1), 2,922 proteins were included.

### Statistical analysis

Figure 1 provides a graphical overview of the analyses performed. All statistical analyses and data cleaning were performed using R version 4.3.2.

We analysed the UKB proteomic data categorised into three protein sets, with differences in the level of statistical evidence required for association detection for each set (described further below):

#### 1) Established biomarkers of neurodegeneration

First, to provide proof-of-concept, we categorised an *a priori* hypothesised set of established biomarkers of neurodegeneration with potential clinical utility (Zetterberg & Schott, 2022). Two proteins were included: glial fibrillary acidic protein (GFAP), a marker of astrocytic activation, and neurofilament light (NEFL; known clinically as NfL), a marker of neurodegeneration and axonal injury. Elevated levels of each are associated with AD, albeit non-specifically (Hansson et al., 2023; Zetterberg & Schott, 2022).

#### 2) Proteins encoded by known genetic risk loci for AD

Second, we categorised an *a priori* hypothesised protein set that overlapped with protein-coding genes identified in the largest, most recent genome-wide association study (GWAS) of AD at the time of analysis (Bellenguez et al., 2022), as summarised in Tables 1 and 2 of that paper. This GWAS included proxy AD cases defined by questionnaire reports of parental dementia; effect sizes for genome-wide significant associations were similar when analyses were restricted to clinically diagnosed AD cases. Of the genes identified, 22 encoded proteins were assayed in UKB.

#### 3) Hypothesis-free comparison of proteomic profiles of APOE ε4 and ε2 carriers with *ε3 homozygotes*

Finally, we analysed all available proteins (*n* = 2,922), including those included in the sets of *a priori* hypothesised proteins, in a hypothesis-free set to examine the proteomic profiles of ε4 or ε2 carriers compared with ε3 homozygotes agnostically.

We applied the same analytical method to three protein sets, differing only in the multiple-testing adjustment. Associations between protein levels and *APOE* ε4 or ε2 carriage (vs ε3 homozygotes) were tested using multivariable linear regression. Analyses were stratified by age (40–50, 50–60, and 60–70 years) to identify age-dependent *APOE* effects on proteomic profiles: 1) to capture associations that may only emerge or become more pronounced in later life as (incipient or manifest) AD emerges for some participants; and 2) to assess how early *APOE*-associated proteomic perturbations can be detected, since AD pathogenesis occurs over decades. To minimise population stratification and account for ancestry-related differences in *APOE*-associated dementia risk (Jackson et al., 2024; Verma et al., 2024), analyses were conducted within homogeneous ancestry groups. Primary analyses were restricted to European ancestry individuals because of the larger sample size and greater statistical power, with additional ancestry-specific analyses performed (see ‘*APOE*-protein associations in AFR and SAS ancestral groups’).

Protein levels were rank-based inverse normal transformed within ancestry and age groups. NPX values below the limit of detection were retained in accordance with Olink recommendations and prior work (Sun et al., 2023). Rank-based inverse normal standardised protein levels were modelled as outcomes with a binary independent variable for ε4 or ε2 carriage. Effect estimates represent the mean standardised difference in relative protein levels (SD units) in ε4 carriers or ε2 carriers relative to ε3 homozygotes. Models were adjusted for age (within strata), sex, proteomic batch, UKB-PPP sub-cohort, and ancestry-specific genetic principal components to control for confounding. Covariate selection was guided by harmonisation with the independent cohorts used in our replication analyses. Because *APOE* genotype is fixed at conception, downstream exposures (e.g., medication use) are unlikely to confound associations, and adjusting for them could induce collider bias; therefore, no post-conception exposures were included in the models. Genetic principal components were derived using the ‘bigsnpr’ package (Privé et al., 2018) following recommended procedures (Privé et al., 2020); full details in Supplementary Methods 2.

Separate regression models were fitted for each protein within each age group; sample sizes varied due to missingness and UKB-PPP quality control (Sun et al., 2023). Multiple-testing correction was not applied to the two *a priori* protein sets, given their established relevance to AD. For hypothesis-free analyses of all other proteins, *P* values were adjusted using the Benjamini-Hochberg procedure, controlling the false discovery rate (FDR) at 5% separately for ε2 and ε4 comparisons.

### Age-related trends in APOE-protein associations

We examined trends in the *APOE*-protein associations across age groups. This analysis was restricted to a subset of prioritised proteins with results generated across age groups. Prioritised proteins were either: 1) from the two *a priori* hypothesised sets; or 2) showing evidence of an association (FDR-adjusted *P* < 0.05) in at least one age group in the hypothesis-free comparison of ε2 or ε4 carriers versus ε3 homozygotes. For hypothesis-free proteins, trends were assessed only within the APOE comparison showing the association. Age trends were tested using meta-regressions (‘rma’ function from the ‘metafor’) package (Viechtbauer, 2010), with mean age per group as the continuous exposure, calculated separately within each analytic sample to account for variability in age distribution across proteins. Again, no multiple testing correction was applied for the two *a priori* hypothesised sets. For proteins identified through the hypothesis-free approach, we adjusted trend *P* values using the Benjamini-Hochberg method (Benjamini & Hochberg, 1995), controlling the FDR at 5%. This adjustment was performed separately within each *APOE* ε2 and ε4 comparison set.

### APOE-protein associations in AFR and SAS ancestral groups

We conducted ancestry-specific analyses in AFR and SAS participants where sample sizes permitted (≥20 ε2 or ε4 carriers per model), but not in East Asian individuals due to insufficient numbers. Analyses followed the same approach as for European ancestry participants, with two exceptions: 1) we did not specifically consider the second list of *a priori* hypothesised proteins, as these had been derived from a GWAS using data from individuals exclusively of European ancestry, and 2) given the smaller sample sizes in these ancestral groups and the exploratory nature of these analyses, we did not apply false discovery rate (FDR) correction, as controlling for Type II error (limiting false negatives) was prioritised over Type I error (production of false positives). Consequently, trends in effect estimates across the three age-stratified groups were assessed for proteins that reached nominal significance (unadjusted *P* < 0.05) in at least one age group, with no adjustment for multiple testing applied in these ancestral groups.

### Pathway enrichment analyses

Pathway enrichment analyses were performed on hypothesis-free results from our primary analytical sample (European ancestry); statistical power was insufficient in other ancestral groups. Analyses were conducted separately for *APOE* ε4- and ε2-associated proteins.

Protein identifiers (UniProt IDs) were mapped to Entrez Gene IDs using the ‘clusterProfiler’ package and the org.Hs.eg.db annotation database. One-to-many mappings were retained, and unmapped proteins were excluded. Gene Ontology (GO) Biological Process gene sets were obtained from the Molecular Signatures Database (MSigDB; collection C5) using ‘msigdbr’. Background gene sets comprised all Entrez-mapped proteins tested in each analysis. Proteins with *P* < 0.05 (FDR corrected) were grouped by age stratum and effect direction. Over-representation analysis (hypergeometric test) was performed using ‘compareCluster()’ with the ‘enrichGO’ method, applying Benjamini-Hochberg correction across all clusters within each *APOE* (ε2 or ε4 carriage) comparison. Gene sets containing fewer than five or more than 500 genes were excluded. To reduce redundancy among enriched GO terms, semantic similarity-based simplification was applied using the ‘Wang method’, retaining one term per similarity cluster (similarity ≥0.7) based on the lowest adjusted *P* value.

### Replication of APOE ε4- and ε2-protein associations in independent cohorts

To replicate key *APOE* ε4- and ε2-associated plasma protein findings (FDR-adjusted *P* < 0.05), including age-related trends, we analysed two independent cohorts (INTERVAL and NSHD) profiled using SomaLogic SomaScan platforms. Ethical approval was granted to INTERVAL by the National Research Ethics Service (11/EE/0538) and to NSHD by the National Research Ethics Service Committee London (14/LO/1173) and Scotland A Research Ethics Committee (14/SS/1009). Full cohort descriptions and replication methods are provided in Supplementary Method 3.

Briefly, genotyping, quality control, imputation, and ancestry assignment were performed centrally by the INTERVAL (Astle et al., 2016) and NSHD (Shireby et al., 2025) study teams. Age-matched subsets of the INTERVAL study (Angelantonio et al., 2017) were used to replicate findings from the youngest (40–50 years, *n* = 705) and middle (50–60 years, *n* = 673) age groups, while the Medical Research Council National Survey of Health and Development (NSHD; 1946 British birth cohort) (Kuh et al., 2016) was used to replicate associations observed in the oldest age group (60–70 years, *n* = 1,645). Plasma protein levels were measured using the SomaScan 4K in INTERVAL (Sun et al., 2018) and the SomaScan 11k in NSHD (Candia et al., 2024; Groves et al., 2025). Coverage of proteins overlapping with those associated with *APOE* ε2 or ε4 in UKB is reported in the Results. Isoform-specific APOE aptamers (APOE2/3/4) on the SomaScan 11K platform were excluded because of poor selectivity (Kirsher et al., 2025); total APOE (non-isoform-specific) was retained across cohorts/platforms. Where multiple SomaScan aptamers mapped to a single UniProt identifier, all were analysed with Bonferroni correction applied (*P* divided by the number of aptamers per UniProt). Replication analyses were restricted to individuals of European ancestry, as sample sizes from other ancestral groups were insufficient for replication.

### Sensitivity analyses

To assess the robustness of our findings, we repeated analyses using z-scored protein levels without rank-based inverse normal transformation and compared effect estimates between transformed and untransformed data. We further compared randomly (*n* = 34,551) and non-randomly selected UKB-PPP participants (*n* = 5,541). In both sensitivity analyses, consistency was evaluated using replication of associations and correlations of effect estimates, stratified by *APOE* carrier status and age group. Full details are provided in Supplementary Method 4.

## Results

### Cohort description

Participant characteristics split by age group and ancestry are reported in Supplementary Table 1. Sample sizes per analysis, including the number of *APOE* ε4 or ε2 carriers, are provided alongside the full results in the Supplementary Tables.

### Associations of *APOE* ε4 and ε2 carriage with established biomarkers of neurodegeneration

Elevated levels of GFAP and NEFL are associated with AD. Among individuals of EUR ancestry (Figure 2A), associations for GFAP and NEFL were directionally consistent with the AD risk conferred by each *APOE* genotype. ε4 carriers had increasingly higher levels of GFAP and NEFL than ε3 homozygotes in the middle and older age groups, with GFAP showing evidence of an age-related trend, while in the youngest group, ε2 carriers showed lower NEFL levels compared to ε3 homozygotes (which was, unexpectedly, not observed in older age groups). There was little evidence of an association between ε2 carriage and GFAP in any age group. In AFR and SAS ancestries (Figure 2B-C), association estimates were less precise, consistent with the smaller sample sizes of these groups. However, among AFR individuals in the oldest age group, ε4 carriage was associated with higher GFAP and NEFL levels. Full results for all ancestries are presented in Supplementary Table 2.

**Figure 2.**
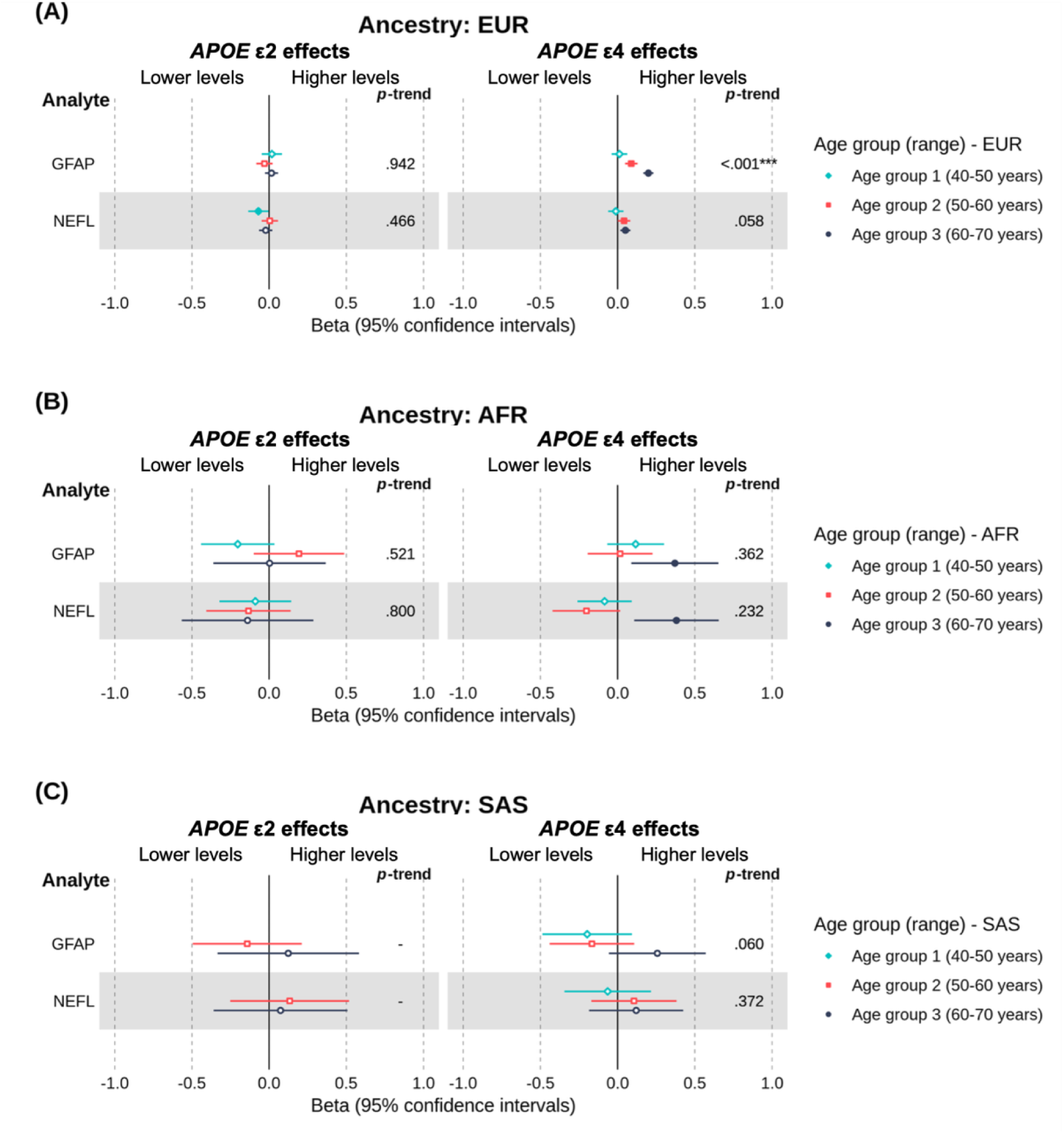
Forest plot of the estimated age- and ancestry-specific effects of *APOE* ε4 and *APOE* ε2 on plasma levels of *a priori* established biomarkers of Alzheimer’s disease, with ε3 homozygotes as the reference group. ***Note.*** EUR = European; AFR = African; SAS = South Asian. This forest plot shows the effect estimates (betas) and 95% confidence intervals for *a priori* established biomarkers of neurodegeneration (see Methods). Positive beta coefficients indicate higher protein levels in ε4 or ε2 carriers, whereas negative beta coefficients indicate lower protein levels, relative to ε3 homozygotes. Betas represent the difference in rank-based inverse normal transformed protein levels (measured in *SD* units relative to the full age group) between carriers and non-carriers, adjusted for covariates. Filled points represent beta estimates with evidence of an association (*P* < 0.05, unadjusted). The left panel shows results for ε4 vs. ε3 homozygotes, whereas the right panel shows the results for the same proteins in ε2 vs. ε3 homozygotes. On the right-hand side of each panel is the *P*-trend value indicating whether there was evidence of an age-related trend in associations across groups (* < 0.05, ** < 0.01, *** < 0.001, unadjusted).

### Proteins encoded by known genetic risk loci for AD

Figure 3 shows the results for proteins encoded by known genetic risk loci for AD (Bellenguez et al., 2022), available in UKB. These analyses were restricted to individuals of EUR ancestry to match the sample in which the risk loci were identified. Both ε2 and ε4 carriage were associated with multiple locus-encoded proteins, with substantial overlap between genotypes. Nine proteins, including CTSB, EPHA1, GRN, IDUA, MME and TREM2, were associated with both ε2 and ε4 carriage (*P* < 0.05, unadjusted, in at least one age group for each allele), typically in opposing directions, whereas several associations were genotype-specific (e.g., BLNK for ε2; CRI, CTSH, LILRB2 and TREML2 for ε4). No evidence of association was observed for a subset of loci in any age group, including ACE and APP (*Ps* ≥ 0.05, unadjusted). Generally, associations were more pronounced at older ages for both ε2 carriers and ε4 carriers (Figure 3). Notably, in the upper age groups, ε2 and ε4 carriage exhibited opposing associations with progranulin (GRN), with levels lowest in ε4. Low progranulin levels play a key role in frontotemporal lobar degeneration (FTLD) (Finch et al., 2009; Wang et al., 2021). Three proteins encoded by known genetic risk loci for AD – IDUA, EPHA1, and MME - showed evidence of age-related trends in *APOE*-protein associations among ε4 carriers (see *P*-trend values in Figure 3). No strong evidence of age-related trends was observed among ε2 carriers, though some tentative evidence was observed for TREM2, where higher levels attenuated toward the null with advancing age. Full results are reported in Supplementary Table 3.

**Figure 3.**
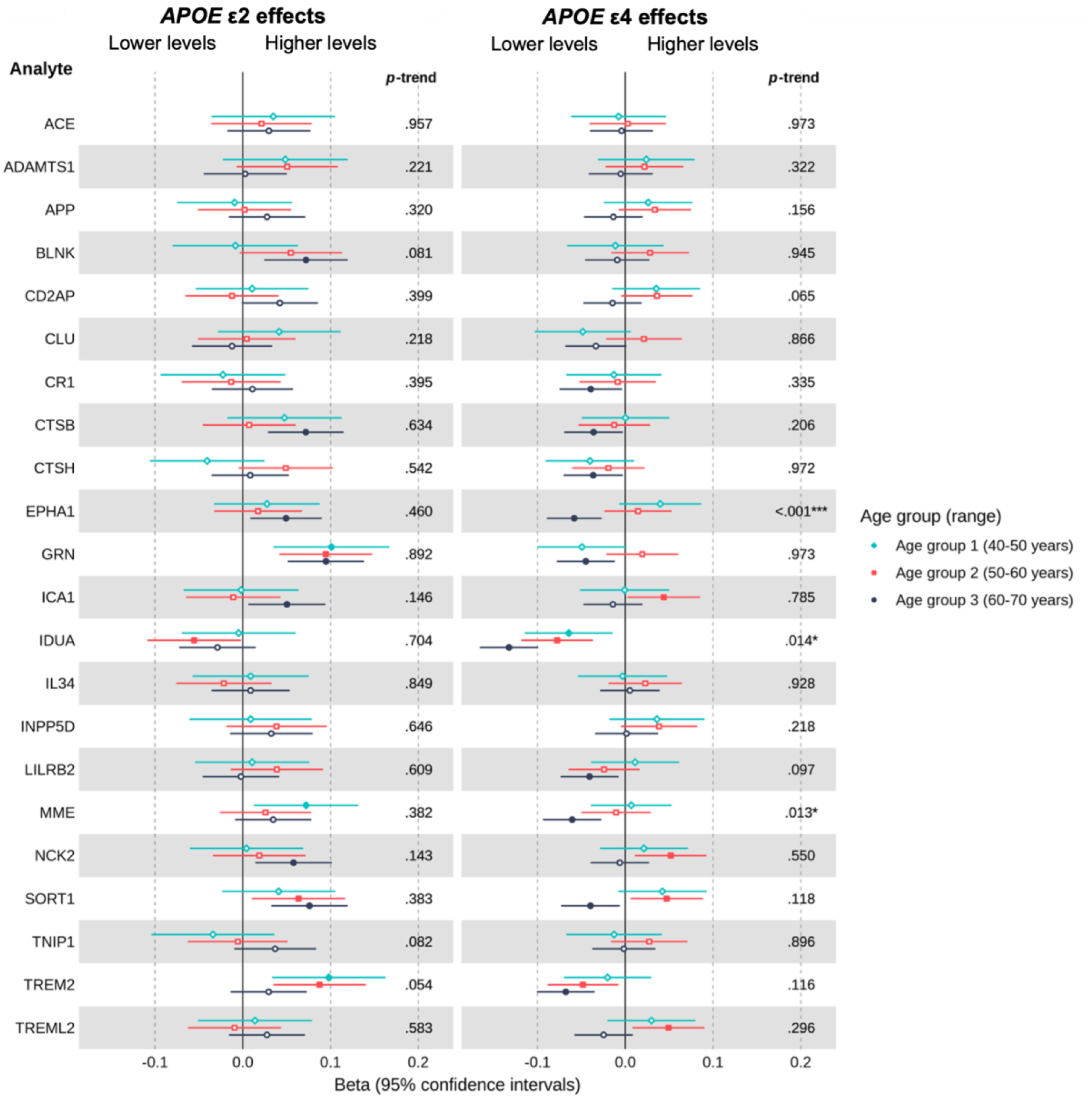
Estimated age-specific effects of *APOE* ε4 and *APOE* ε2 on plasma levels of proteins encoded by known genetic risk loci for Alzheimer’s disease in European ancestry individuals, with ε3 homozygotes as the reference group. ***Note*.** Betas represent the difference in rank-based inverse normal transformed protein levels (measured in *SD* units relative to the full age group) between ε4 or ε2 carriers and ε3 homozygotes, adjusted for covariates. Positive beta coefficients indicate higher protein levels in ε4 or ε2 carriers, whereas negative beta coefficients indicate lower protein levels, relative to ε3 homozygotes. Filled points represent beta estimates with *P* < 0.05 (unadjusted). The left panel shows results for ε2 vs. ε3 homozygotes, whereas the right panel shows the results for the same proteins in ε4 vs. ε3 homozygotes. On the right-hand side of each panel is the *P-*trend value indicating whether there was evidence of an age-related trend in associations across groups (* < 0.05, ** < 0.01, *** < 0.001, unadjusted).

### Hypothesis-free comparison of proteomic profiles of *APOE* ε4 and ε2 carriers with ε3 homozygotes

In our hypothesis-free analyses, we observed distinct proteomic profiles for *APOE* ε4 and ε2 carriers. Among individuals of EUR ancestry, 480 proteins were associated with ε4 carriage and 351 with ε2 carriage (FDR-adjusted *P* < 0.05 in at least one age group), with 130 proteins shared between genotypes. A subset showed particularly robust associations across age groups (FDR-adjusted *P* < 0.001 in ≥2 groups), with 28 ε4- and 25 ε2-associated proteins highlighted in Figure 4 and Figure 5, respectively. Several proteins, including APOE, PLA2G7, BRK1 and ANGPTL3, demonstrated opposing effects in ε4 versus ε2 carriers, whereas AGRP and MENT were directionally concordant, with higher levels in both ε4 and ε2 carriers compared to the ε3 homozygote reference group. In contrast, several proteins showed evidence of associations exclusively in either ε2 or ε4 carriers (versus ε3 homozygotes). ε2, but not ε4, was associated with levels of APOC1, BCAM, BGLAP, CCL27, CCN1, DEFB4A_DEFB4B, FGFBP1, FGFBP2, HMOX1, IFNGR2, ITGA2, LDLR, MXRA8, NPY, PDGFC, PLA2G10, PVR, S100P, and TFPI. Whereas ε4, but not ε2, was associated with levels of AGR3, APOA2, APOF, BPIFB2, CES1, CLEC4A, CPQ, CREG1, CSNK2A1, CTSZ, ERN1, GFAP, HMOX2, IL32, MAN2B2, NAAA, PALM, RP2, SMPD1, SMPDL3A, SNAP25, VNN1. Considering proteins most robustly associated with ε2 or ε4 carriership (Figures 4 & 5), some evidence of age-related trends was observed, though effect sizes were generally relatively consistent across age groups, with greater variation between proteins than across age groups for a given protein. More broadly, across all available proteins, there was evidence for age-related trends in *APOE*-protein associations for 25 proteins (FDR-adjusted *Ps* < 0.05). All 25 trends were observed within ε4 carrier comparisons with ε3 homozygotes. There was little evidence for age trends where ε2 carriership was associated with the protein in at least one group. Full ε4 and ε2 results for EUR individuals are provided in Supplementary Tables 4 and 5, respectively.

**Figure 4.**
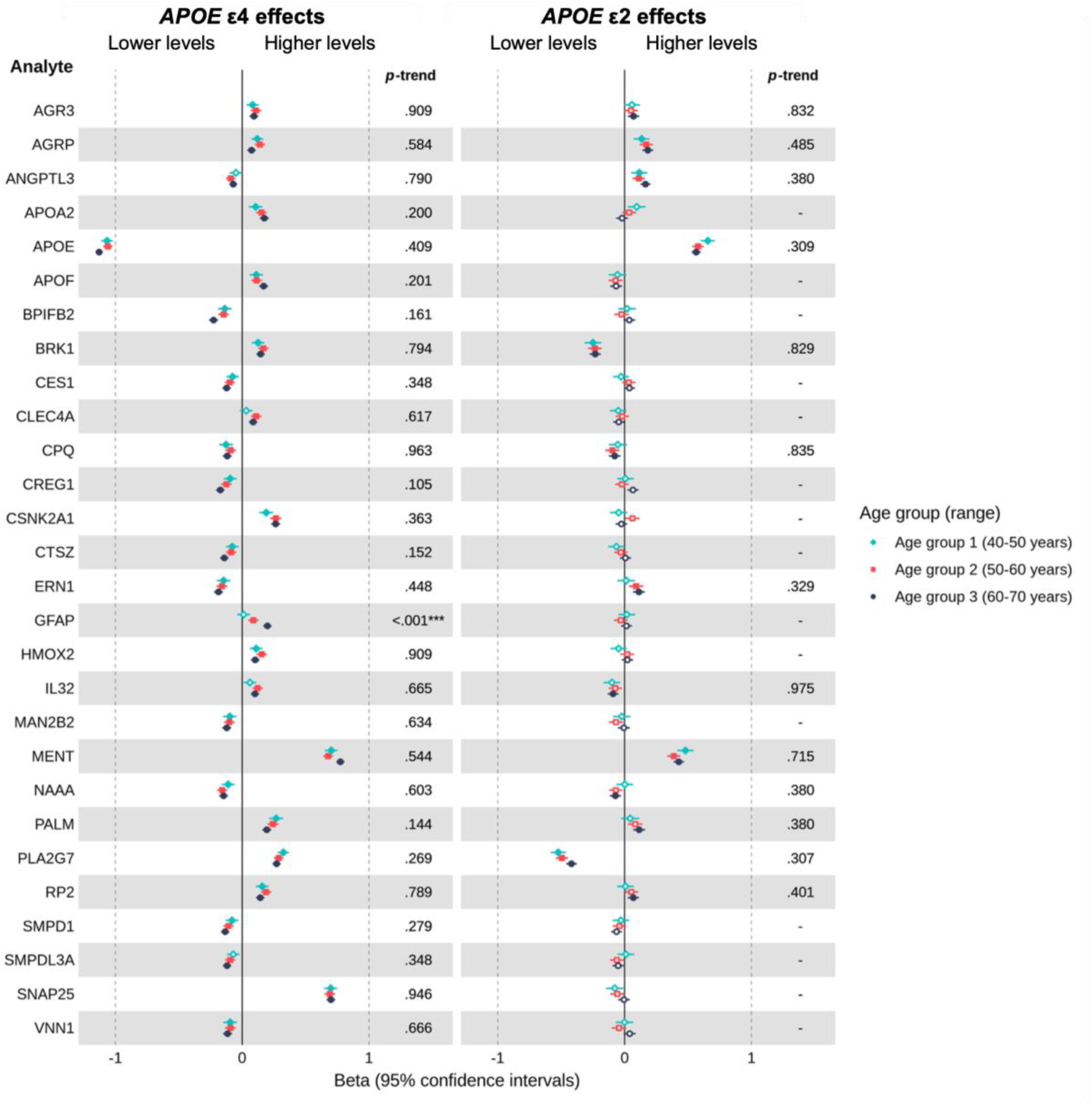
Estimated age-specific effects of *APOE* ε4 (and *APOE* ε2 for comparison) on plasma levels of proteins consistently associated with *APOE* ε4 carrier status in individuals of European ancestry, relative to *APOE* ε3 homozygotes. ***Note.*** Betas represent the difference in rank-based inverse normal transformed protein levels (measured in *SD* units relative to the full age group) between ε4 or ε2 carriers and ε3 homozygotes, adjusted for covariates. Positive beta coefficients indicate higher protein levels in ε4 or ε2 carriers, whereas negative beta coefficients indicate lower protein levels, relative to ε3 homozygotes. Filled points represent beta estimates with a *P* < 0.05 (FDR-adjusted). The left panel shows results for ε4 vs. ε3 homozygotes, whereas the right panel shows the results for the same proteins in ε2 vs. ε3 homozygotes. On the right-hand side of each panel is the FDR-adjusted *P*-trend value indicating whether there was evidence of an age-related trend in associations across groups (* < 0.05, ** < 0.01, *** < 0.001, unadjusted). Trends were only estimated for proteins showing evidence of an association (FDR-adjusted *P* < 0.05) in at least one age group, for the corresponding comparison of ε4 or ε2 carriers versus ε3 homozygotes.

**Figure 5.**
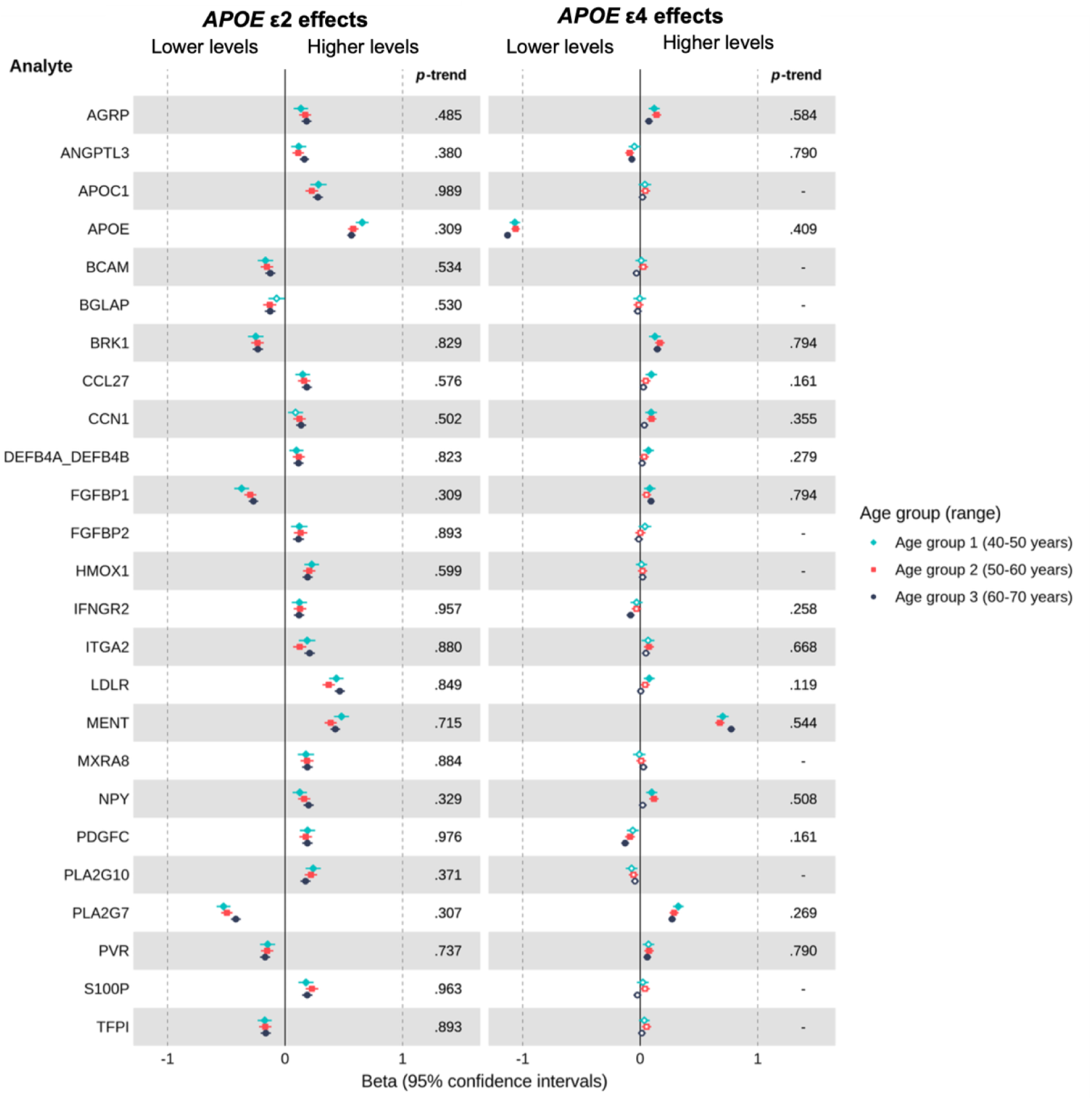
Estimated age-specific effects of *APOE* ε2 (and *APOE* ε4 for comparison) on plasma levels of proteins consistently associated with *APOE* ε2 carrier status in individuals of European ancestry, relative to *APOE* ε3 homozygotes. ***Note*.** Betas represent the difference in rank-based inverse normal transformed protein levels (measured in *SD* units relative to the full age group) between ε4 or ε2 carriers and ε3 homozygotes, adjusted for covariates. Positive beta coefficients indicate higher protein levels in ε4 or ε2 carriers, whereas negative beta coefficients indicate lower protein levels, relative to ε3 homozygotes. Filled points represent beta estimates with *P* <0.05 (FDR-adjusted). The left panel shows results for ε2 vs. ε3 homozygotes, whereas the right panel shows the results for the same proteins in ε4 vs. ε3 homozygotes. On the right-hand side of each panel is the FDR-adjusted *P*-trend value indicating whether there was evidence of an age-related trend in associations across groups (< 0.05, ** < 0.01, *** < 0.001, unadjusted). Trends were only estimated for proteins showing evidence of an association (FDR-adjusted *P* <0.05) in at least one age group, for the corresponding comparison of ε4 or ε2 carriers versus ε3 homozygotes.

In individuals of AFR and SAS ancestry, we also identified many proteins associated with ε2 or ε4 carriage. In AFR participants, 470 proteins showed evidence of association with ε4 carriage and 284 with ε2 carriage, with partial overlap with EUR findings (31.9% for ε4; 25.7% for ε2), and 51 ε4- and 43 ε2-associated proteins exhibited age-related trends. In SAS participants, 112 proteins were associated with ε4 and 92 with ε2 carriage, with 30.2% (ε4) and 28.0% (ε2) overlapping EUR results; age-related trends were observed for 49 ε4-associated proteins, while ε2 analyses were limited by insufficient numbers in the youngest age group (fewer than 20 ε2 carriers). In both AFR and SAS individuals, APOE, MENT, and SNAP25 were robustly associated with ε4 carriage, with concordant effect directions and magnitudes (negative for APOE; positive for MENT and SNAP25). Notably, in individuals of AFR ancestry, ε2 carriage was also robustly associated with higher APOE levels (unadjusted *P* < 0.05 in at least one age group). Full replication results are reported in Supplementary Tables 6-9; Supplementary Figures 1-2. These findings should be interpreted cautiously, given smaller sample sizes and the use of unadjusted *P* values (see Methods).

#### Pathway enrichment analyses

Pathway enrichment analyses demonstrated distinct, age-dependent biological processes associated with *APOE* ε4 and ε2 carriage relative to ε3 homozygotes among individuals of European ancestry. ε4 carriage was characterised by enrichment of lipid and lipoprotein metabolic processes in younger participants (40-50 years), with the strongest signals observed in the oldest age group for immune response, metabolism, lipid storage, cell movement/development pathways (60-70 years) (Supplementary Table 10). In contrast, ε2 carriage was primarily associated with enrichment of lipid and lipoprotein metabolic pathways in younger and middle-aged groups (40-50 and 50-60 years), but at older ages (60-70 years), enrichment was observed for cell-signalling and RNA-related pathways (Supplementary Table 11). Fewer enriched pathways were detected among ε2 carriers in the oldest age group, suggesting attenuation or increased heterogeneity of ε2-associated biological processes with advancing age.

#### Replication of APOE ε4- and ε2-protein associations in independent cohort SomaLogic data

To validate *APOE* ε4- and ε2-associated effects on plasma proteins identified in UKB (FDR-adjusted *P* < 0.05 or *P* < 0.05 for *a priori* hypothesised proteins), we performed approximately age-matched analyses in two independent cohorts profiled using SomaLogic assays: INTERVAL (SomaScan 4K v3.0) and NSHD (SomaScan 11K v5.0). Replication was defined as nominal evidence of association (*P* < 0.05) with directionally concordant effects relative to UKB. Cohort characteristics, sample sizes, and replication statistics are in Supplementary Tables 12–14, with further description in Supplementary Result 1.

In the 40-50-year age group, 59.4% of UKB-identified proteins were available in INTERVAL, of which four ε4-associated proteins (LDLR, MENT, NAAA and PLA2G7) and one ε2-associated protein (PLA2G7) replicated. In the 50-60-year age group, 55.6% were available, with six ε4- and ten ε2-associated proteins. Platform coverage was most extensive for the oldest age group (60-70 years in UKB; 60-65 years in NSHD), where 92.9% of proteins were available, with 25 ε4- and 26 ε2-associated proteins replicated. Replicated ε4-associated proteins included several showing robust associations in UKB (e.g., APOE, ANGPTL3, MENT and PLA2G7), whereas ε2-associated proteins encompassed proteins encoded by established Alzheimer’s disease risk loci (CTSB and GRN) and others showing robust association in UKB (e.g., MENT and PLA2G7).

A few proteins showed cross-platform discordance in age-matched results between UKB and the replication cohorts. For example, NEFL in ε4 carriers displayed opposite effects to UKB (i.e., lower levels), consistent with previous reports of SomaScan-specific inverse associations with Alzheimer’s risk (Budelier et al., 2022; Frick et al., 2024; Leckey & Zetterberg, 2022); NEFL was not measured in INTERVAL. SNAP25 and GRN also showed inconsistent effects in older ε4 carriers (opposite directions of effect to UKB), potentially reflecting assay-specific differences.

Limited replication of age-related trends was observed. Among the 14 of 25 (56%) ε4-associated proteins showing age trends in UKB that were measured in replication cohorts, none reached nominal significance (*Ps* > 0.05). No age-dependent protein trends were identified among ε2 carriers in UKB, precluding replication analyses for this group.

### Sensitivity analyses

Sensitivity analyses confirmed the general robustness of our findings, with and without rank-based inverse normal transformation (Supplementary Tables 16-18), and across random and non-random UKB-PPP sub-cohorts when restricting to proteins associated with *APOE* ε4 or ε2 in at least one age group in the non-random sample (Supplementary Tables 19-23). A more detailed summary is provided in Supplementary Result 2.

## Discussion

In this large-scale plasma proteomic analysis stratified by age, we demonstrate that *APOE* ε4 and ε2 carriage are associated with distinct proteomic profiles, detectable at least from midlife. These proteins included established biomarkers of AD, proteins encoded by known AD genetic risk loci, and novel and established associations among a broad set of proteins identified through a hypothesis-free, proteome-wide approach. Together, our findings highlight widespread, partly age-dependent alterations in circulating proteins associated with *APOE* variation, some of which may be related to preclinical AD (either as mediating cause or consequence of disease pathogenesis).

As anticipated, *APOE* variation was associated with two well-established biomarkers of neurodegeneration (Hansson et al., 2023; Zetterberg & Schott, 2022). Elevated plasma levels of GFAP and NEFL have been linked to increased 10-year risk of incident all-cause dementia (Guo et al., 2024). The presence of a GFAP effect in ε4 but not ε2 carriers may reflect the greater Aβ accumulation and earlier AD onset observed in ε4 carriers (Christensen et al., 2010; Polsinelli et al., 2023; Yamazaki et al., 2019). Differences in GFAP levels between ε2 carriers and ε3 homozygotes may emerge later in life, beyond the age range studied here, in line with epidemiological evidence that separation in dementia risk between ε2 and ε3 occurs later than that observed for ε4 versus ε3 (Reiman et al., 2020). Alternatively, ε2 and ε3 carriers exhibit different molecular correlates of disease risk to ε4 carriers, rather than delayed manifestation alone, in line with prior work demonstrating *APOE* ε4-dependent-and-independent proteomic signatures of incident Alzheimer’s disease (Frick et al., 2024).

ε4 carriers showed patterns of markedly lower levels of several proteins encoded by AD genetic risk loci in the oldest group relative to ε3 homozygotes (e.g., IDUA, MME, GRN, SORT1, TREM2); notably, SORT1 reversed direction across age, from higher levels in midlife (50–60 years) to lower levels in later life (60–70 years). Mendelian randomisation studies in individuals of European ancestry provide evidence for causal associations between higher circulating progranulin (GRN) and TREM2 (triggering receptor expressed on myeloid cells 2) levels and reduced Alzheimer’s disease risk (Liu et al., 2024; Zhan et al., 2025), consistent with suggestions of neuroprotection (for GRN) and enhanced microglial clearance (for TREM2) (Rhinn et al., 2022; L. Zhang et al., 2025). Together, these findings may suggest potential age-related attenuation of neuroprotective mechanisms in ε4 carriers during this age range. More broadly, accumulating evidence suggests an interplay between *APOE* genotype and age in shaping both circulating and cerebrospinal fluid (CSF) molecular profiles, including metabolites and proteins associated with Alzheimer’s disease and related pathology (Amin et al., 2025; Compton et al., 2024; Wesenhagen et al., 2022).

In our hypothesis-free, proteome-wide analyses, several proteins showed strong associations with *APOE* variation across multiple ages, ancestral populations, and were replicated in independent cohorts (e.g., APOE, MENT, PLA2G7). The gradient in circulating APOE levels from higher in ε2, intermediate in ε3, and lower in ε4 has been previously established, but does not appear to be reflected by APOE levels in the central nervous system (Lawler et al., 2023). Whether circulating levels of APOE are specifically of aetiological relevance to AD, or confounded by APOE genotype, is unclear. Nonetheless, other proteins for which there are gradients in concentrations across ε2, ε3, and ε4 alleles (e.g. PLA2G7; platelet-activating factor acetylhydrolase), or for which there is the same direction of effects for ε2 and ε4 (most notably for MENT; methylated in normal thymocytes protein), may hold mechanistic insights into how *APOE* affects AD risk. For example, a recent study of UK Biobank participants aged 50 and older found that PLA2G7 was not a direct mediator of Alzheimer’s disease risk, but it showed a weak mediated interaction with a polygenic risk score that included *APOE* variation, suggesting a subtle modulatory role (Beydoun et al., 2025)

Many of the proteins identified here have been linked to multiple neurodegenerative diseases beyond AD, including GRN (Rhinn et al., 2022), SNAP25 (synaptosomal-associated protein of 25 kDa) (C. Zhang et al., 2025), and TREM2 (Carmona et al., 2018), highlighting potentially shared *APOE*-related molecular pathways to different outcomes. Notably, effects on GRN levels - a biomarker decreased in carriers of GRN mutations causing frontotemporal dementia (FTD) (Finch et al., 2009; Wang et al., 2021) - were more pronounced and observed at younger ages in ε2 compared with ε4 carriers. In contrast, among the oldest individuals, circulating progranulin levels showed a decreasing gradient across ε2, ε3, and ε4, a pattern that, to our knowledge, has not been previously reported. However, this finding was only partly replicated in an age-matched independent cohort (NSHD) profiled using the SomaScan platform, where both ε2 and ε4 were associated with higher GRN levels. Therefore, further validation is required, as differences in proteomic platforms may partly underlie this discrepancy, and others (e.g., NEFL, SNAP25). Overall, our results align with recent evidence of a conserved *APOE* ε4-associated pro-inflammatory immune signature across the brain, CSF, and plasma, across neurodegenerative diseases and non-impaired individuals, suggesting a shared vulnerability that precedes clinical onset (Shvetcov et al., 2025). However, *APOE* exhibits broad pleiotropic effects across diverse health outcomes (e.g., type 2 diabetes, liver disease, ischaemic heart disease), with evidence suggesting both protective and harmful effects of ε4 and ε2 varying dependent on disease context (Lumsden et al., 2020). Thus, some ε4-associated proteins identified here may not necessarily reflect pathways increasing neurodegeneration risk, and conversely, some ε2-associated proteins may not be protective. Together, our results add to growing evidence suggesting a broader role of *APOE* variation across neurodegenerative diseases, warranting further investigation with potential to inform disease-specific biomarkers and therapeutic strategies targeting shared pathways.

### Strengths and limitations

To our knowledge, this study provides the largest age-stratified analysis to date of plasma proteomic profiles associated with *APOE* ε4 and ε2 carriage (*N* = 42,642). Key strengths include its broad proteomic coverage (Olink Explore 3072), stratification across adults aged approximately 40-70 years, inclusion of multiple ancestral groups, and replication of some findings in independent cohorts using a distinct proteomic platform (SomaScan). However, several limitations should be considered.

First, platform-related differences in protein coverage limited our ability to replicate key proteins, particularly in the youngest and middle age groups where coverage was most limited. However, such data are limited due to the high cost of high-throughput proteomic profiling. Second, the cross-sectional design precludes inference about within-individual ageing trajectories, and observed age-related trends may partly reflect cohort effects arising from differences in life course exposures which may modify how *APOE* variation affects the proteome (e.g., healthcare or socioeconomic factors). Longitudinal studies with repeated measures within individuals would help replicate age-related trends. Third, selection bias likely influenced our results, as UKB participants tend to be healthier, wealthier, and more educated than the general population, limiting generalisability (Schoeler et al., 2023). More specifically, survivor bias could have impacted age-related trends, particularly for *APOE* ε4, which is linked to premature mortality among individuals of European ancestry (Wolters et al., 2019). However, the similar distribution of *APOE* genotypes across age groups suggests that any such bias is largely independent of genotype. Similarly, it is challenging to determine whether observed *APOE*-associated proteomic differences reflect primary biological effects of genotype or secondary consequences of other factors, such as medication initiation. For example, *APOE* genotype influences cardiometabolic risk traits across diverse populations (Verma et al., 2024), which can prompt the initiation of pharmacotherapies such as statins and antihypertensives that may affect circulating protein levels (Bohn et al., 2023; Enroth et al., 2018). Finally, smaller sample sizes limited statistical power in analyses of AFR and SAS ancestry groups and precluded analyses in East Asian individuals entirely. Future studies in larger, ancestrally diverse cohorts could clarify potential ancestry-specific effects of *APOE* variation on the plasma proteome, including potential gene-by-environment or local ancestry interactions, thereby improving understanding of AD pathophysiology and supporting equitable biomarker and therapeutic development.

### Future directions

Further work is needed to establish the aetiological relevance of these proteins to AD and other neurodegenerative diseases, and to determine whether plasma and CSF show overlapping or distinct *APOE*- and age-dependent effects. Irrespective of causality, plasma proteomics may offer minimally invasive, clinically informative signals that complement CSF measures.

## Supporting information

Supplementary materials: Methods, Results and Figures

## Acknowledgments

We are grateful to the study participants and teams of the INTERVAL, NSHD and UKB cohorts for their invaluable help toward the generation and curation of data used in this project. This research has been conducted using the UK Biobank Resource under Application Number 12113. We thank the Alzheimer’s Society monitors - Angela Clayton-Turner, Bernadette Roden, and Mary Sherrington - for their guidance and feedback on our research and related outputs. The authors acknowledge the use of the UCL Data Safe Haven and Myriad High Performance Computing Facility (Myriad@UCL), and associated support services, in the completion of this work.

## Funding sources

This research was supported by funding from Alzheimer’s Society (grant number 633, AS-PG-23-021). DMW is supported by an Alzheimer’s Research UK Senior Fellowship (ARUK-SRF2023B-008). ELA is supported by a UKRI Future Leaders Fellowship (MR/W011581/1). Proteomic work in NSHD was supported by the Phil and Penny Knight Initiative for Brain Resilience, the Alzheimer’s Association (T.W.-C.), the Simons Foundation (T.W.-C.), and the National Institute on Aging (AG072255, T.W.-C), the Milky Way Research Foundation (T.W.-C.). JMS is a National Institute for Health Research (NIHR) Senior Investigator and acknowledges the support of the NIHR University College London Hospitals Biomedical Research Centre and the UCL Centre of Research Excellence, an initiative funded by British Heart Foundation (RE/24/130013). This work is supported by the UK Dementia Research Institute through UK DRI Ltd, principally funded by the Medical Research Council and grant funding from Alzheimer’s Research UK, LifeArc, Brain Research UK, Weston Brain Institute, Wolfson Foundation, and Alzheimer’s Association (SG-666374-UK BIRTH COHORT). J.W.G. is supported by an Alzheimer’s Research UK (ARUK) Clinical Research Training Fellowship (ARUK-CRTF2023B-001). Participants in the INTERVAL randomised controlled trial were recruited with the active collaboration of NHS Blood and Transplant England (www.nhsbt.nhs.uk), which has supported field work and other elements of the trial. DNA extraction and genotyping were co-funded by the National Institute for Health and Care Research (NIHR), the NIHR BioResource (http://bioresource.nihr.ac.uk) and the NIHR Cambridge Biomedical Research Centre (BRC-1215-20014) [*]. The academic coordinating centre for INTERVAL was supported by core funding from the: NIHR Blood and Transplant Research Unit (BTRU) in Donor Health and Genomics (NIHR BTRU-2014-10024), NIHR BTRU in Donor Health and Behaviour (NIHR203337), UK Medical Research Council (MR/L003120/1), British Heart Foundation (SP/09/002; RG/13/13/30194; RG/18/13/33946) and NIHR Cambridge BRC (BRC-1215-20014; NIHR203312) [*]. A complete list of the investigators and contributors to the INTERVAL trial is provided in reference (Angelantonio et al., 2017). The academic coordinating centre would like to thank blood donor centre staff and blood donors for participating in the INTERVAL trial. SomaLogic assays were funded by Merck and the NIHR Cambridge Biomedical Research Centre (NIHR203312). [*] The views expressed are those of the authors and not necessarily those of the NIHR or the Department of Health and Social Care.

## Data availability

Data are available from the UK Biobank upon application (https://www.ukbiobank.ac.uk). Bona fide researchers can apply to access the NSHD data via a standard application procedure (further details available at https://skylark.ucl.ac.uk/NSHD/access/). The INTERVAL study data used in this paper are available to bona fide researchers from. A GitLab repository containing analysis code will be made available at: https://gitlab.com/aepacker/apoe_plasma_proteomics/.

## Conflicts of interest statement

T.W.-C. is a co-founder and scientific advisor of Vero Biosciences and Teal Omics and holds equity stakes in these companies. J.M.S. has received research funding and PET tracer from AVID Radiopharmaceuticals (a wholly owned subsidiary of Eli Lilly) and Alliance Medical; has consulted for Roche, Eli Lilly, Biogen, MSD, GE Healthcare and Alamar Biosciences; and received royalties from Oxford University Press and Henry Stewart Talks. He is Chief Medical Officer for Alzheimer’s Research UK.

## Notes

### Author Declarations

UK Biobank data has ethical approval from the North-West Multi-centre Research Ethics Committee (REC reference numbers 11/NW/0382 and 16/NW/0274). Ethical approval was granted to INTERVAL by the National Research Ethics Service (11/EE/0538) and to NSHD by the National Research Ethics Service Committee London (14/LO/1173) and Scotland A Research Ethics Committee (14/SS/1009).

## References

Amin, N., Liu, J., Sproviero, W., Arnold, M., Batra, R., Bonnechere, B., Chiou, Y.-J., Fernandes, M., Krumsiek, J., Newby, D., Nho, K., Kim, J. P., Saykin, A. J., Shi, L., Winchester, L. M., Yang, Y., Nevado-Holgado, A. J., Kastenmüller, G., Kaddurah-Daouk, R., & van Duijn, C. M. (2025). Interplay between age, APOE Ɛ4 and the metabolome in plasma and brain in Alzheimer’s disease. Translational Psychiatry, 15, 460. 10.1038/s41398-025-03625-8

Angelantonio, E. D., Thompson, S. G., Kaptoge, S., Moore, C., Walker, M., Armitage, J., Ouwehand, W. H., Roberts, D. J., Danesh, J., Angelantonio, E. D., Thompson, S. G., Kaptoge, S., Moore, C., Walker, M., Armitage, J., Ouwehand, W. H., Roberts, D. J., Danesh, J., Armitage, J., … Roberts, D. J. (2017). Efficiency and safety of varying the frequency of whole blood donation (INTERVAL): A randomised trial of 45 000 donors. The Lancet, 390(10110), 2360–2371. 10.1016/S0140-6736(17)31928-1

Astle, W. J., Elding, H., Jiang, T., Allen, D., Ruklisa, D., Mann, A. L., Mead, D., Bouman, H., Riveros-Mckay, F., Kostadima, M. A., Lambourne, J. J., Sivapalaratnam, S., Downes, K., Kundu, K., Bomba, L., Berentsen, K., Bradley, J. R., Daugherty, L. C., Delaneau, O., … Soranzo, N. (2016). The Allelic Landscape of Human Blood Cell Trait Variation and Links to Common Complex Disease. Cell, 167(5), 1415–1429.e19. 10.1016/j.cell.2016.10.042

Auton, A., Abecasis, G. R., Altshuler, D. M., Durbin, R. M., Abecasis, G. R., Bentley, D. R., Chakravarti, A., Clark, A. G., Donnelly, P., Eichler, E. E., Flicek, P., Gabriel, S. B., Gibbs, R. A., Green, E. D., Hurles, M. E., Knoppers, B. M., Korbel, J. O., Lander, E. S., Lee, C., … National Eye Institute, N. (2015). A global reference for human genetic variation. Nature, 526(7571), 68–74. 10.1038/nature15393

Bellenguez, C., Küçükali, F., Jansen, I. E., Kleineidam, L., Moreno-Grau, S., Amin, N., Naj, A. C., Campos-Martin, R., Grenier-Boley, B., Andrade, V., Holmans, P. A., Boland, A., Damotte, V., van der Lee, S. J., Costa, M. R., Kuulasmaa, T., Yang, Q., de Rojas, I., Bis, J. C., … Lambert, J.-C. (2022). New insights into the genetic etiology of Alzheimer’s disease and related dementias. Nature Genetics, 54(4), 412–436. 10.1038/s41588-022-01024-z

Benedet, A. L., Milà-Alomà, M., Vrillon, A., Ashton, N. J., Pascoal, T. A., Lussier, F., Karikari, T. K., Hourregue, C., Cognat, E., Dumurgier, J., Stevenson, J., Rahmouni, N., Pallen, V., Poltronetti, N. M., Salvadó, G., Shekari, M., Operto, G., Gispert, J. D., Minguillon, C., … Translational Biomarkers in Aging and Dementia (TRIAD) study, A. and F. (ALFA) study, and BioCogBank Paris Lariboisière cohort. (2021). Differences Between Plasma and Cerebrospinal Fluid Glial Fibrillary Acidic Protein Levels Across the Alzheimer Disease Continuum. JAMA Neurology, 78(12), 1471–1483. 10.1001/jamaneurol.2021.3671

Benjamini, Y., & Hochberg, Y. (1995). Controlling the False Discovery Rate: A Practical and Powerful Approach to Multiple Testing. Journal of the Royal Statistical Society: Series B (Methodological), 57(1), 289–300. 10.1111/j.2517-6161.1995.tb02031.x

Beydoun, M. A., Beydoun, H. A., Li, Z., Hu, Y.-H., Noren Hooten, N., Ding, J., Hossain, S., Maino Vieytes, C. A., Launer, L. J., Evans, M. K., & Zonderman, A. B. (2025). Alzheimer’s Disease polygenic risk, the plasma proteome, and dementia incidence among UK older adults. GeroScience, 47(2), 2507–2523. 10.1007/s11357-024-01413-8

Bohn, B., Lutsey, P. L., Tang, W., Pankow, J. S., Norby, F. L., Yu, B., Ballantyne, C. M., Whitsel, E. A., Matsushita, K., & Demmer, R. T. (2023). A Proteomic Approach for Investigating the Pleiotropic Effects of Statins in the Atherosclerosis Risk in Communities (ARIC) Study. Journal of Proteomics, 272, 104788. 10.1016/j.jprot.2022.104788

Bridel, C., van Wieringen, W. N., Zetterberg, H., Tijms, B. M., Teunissen, C. E., & and the NFL Group. (2019). Diagnostic Value of Cerebrospinal Fluid Neurofilament Light Protein in Neurology: A Systematic Review and Meta-analysis. JAMA Neurology, 76(9), 1035–1048. 10.1001/jamaneurol.2019.1534

Budelier, M. M., He, Y., Barthelemy, N. R., Jiang, H., Li, Y., Park, E., Henson, R. L., Schindler, S. E., Holtzman, D. M., & Bateman, R. J. (2022). A map of neurofilament light chain species in brain and cerebrospinal fluid and alterations in Alzheimer’s disease. Brain Communications, 4(2). 10.1093/braincomms/fcac045

Bycroft, C., Freeman, C., Petkova, D., Band, G., Elliott, L. T., Sharp, K., Motyer, A., Vukcevic, D., Delaneau, O., O’Connell, J., Cortes, A., Welsh, S., Young, A., Effingham, M., McVean, G., Leslie, S., Allen, N., Donnelly, P., & Marchini, J. (2018). The UK Biobank resource with deep phenotyping and genomic data. Nature, 562(7726), 203–209. 10.1038/s41586-018-0579-z

Candia, J., Fantoni, G., Delgado-Peraza, F., Shehadeh, N., Tanaka, T., Moaddel, R., Walker, K. A., & Ferrucci, L. (2024). Variability of 7K and 11K SomaScan Plasma Proteomics Assays. Journal of Proteome Research, 23(12), 5531–5539. 10.1021/acs.jproteome.4c00667

Carmona, S., Zahs, K., Wu, E., Dakin, K., Bras, J., & Guerreiro, R. (2018). The role of *TREM2* in Alzheimer’s disease and other neurodegenerative disorders. The Lancet Neurology, 17(8), 721–730. 10.1016/S1474-4422(18)30232-1

Christensen, D. Z., Schneider-Axmann, T., Lucassen, P. J., Bayer, T. A., & Wirths, O. (2010). Accumulation of intraneuronal Aβ correlates with ApoE4 genotype. Acta Neuropathologica, 119(5), 555–566. 10.1007/s00401-010-0666-1

Compton, H., Smith, M. L., Bull, C., Korologou-Linden, R., Ben-Shlomo, Y., Bell, J. A., Williams, D. M., & Anderson, E. L. (2024). Life course plasma metabolomic signatures of genetic liability to Alzheimer’s disease. Scientific Reports, 14(1), 3896. 10.1038/s41598-024-54569-w

Cui, M., Cheng, C., & Zhang, L. (2022). High-throughput proteomics: A methodological mini-review. Laboratory Investigation, 102(11), 1170–1181. 10.1038/s41374-022-00830-7

Enroth, S., Maturi, V., Berggrund, M., Enroth, S. B., Moustakas, A., Johansson, Å., & Gyllensten, U. (2018). Systemic and specific effects of antihypertensive and lipid-lowering medication on plasma protein biomarkers for cardiovascular diseases. Scientific Reports, 8, 5531. 10.1038/s41598-018-23860-y

Finch, N., Baker, M., Crook, R., Swanson, K., Kuntz, K., Surtees, R., Bisceglio, G., Rovelet-Lecrux, A., Boeve, B., Petersen, R. C., Dickson, D. W., Younkin, S. G., Deramecourt, V., Crook, J., Graff-Radford, N. R., & Rademakers, R. (2009). Plasma progranulin levels predict progranulin mutation status in frontotemporal dementia patients and asymptomatic family members. Brain, 132(3), 583–591. 10.1093/brain/awn352

Frick, E. A., Emilsson, V., Jonmundsson, T., Steindorsdottir, A. E., Johnson, E. C. B., Puerta, R., Dammer, E. B., Shantaraman, A., Cano, A., Boada, M., Valero, S., García-González, P., Gudmundsson, E. F., Gudjonsson, A., Pitts, R., Qiu, X., Finkel, N., Loureiro, J. J., Orth, A. P., … Gudnason, V. (2024). Serum proteomics reveal APOE-ε4-dependent and APOE-ε4-independent protein signatures in Alzheimer’s disease. Nature Aging, 4(10), 1446–1464. 10.1038/s43587-024-00693-1

Frisoni, G. B., Fox, N. C., Jack, C. R., Scheltens, P., & Thompson, P. M. (2010). The clinical use of structural MRI in Alzheimer disease. Nature Reviews. Neurology, 6(2), 67–77. 10.1038/nrneurol.2009.215

Fry, A., Littlejohns, T. J., Sudlow, C., Doherty, N., Adamska, L., Sprosen, T., Collins, R., & Allen, N. E. (2017). Comparison of Sociodemographic and Health-Related Characteristics of UK Biobank Participants With Those of the General Population. American Journal of Epidemiology, 186(9), 1026–1034. 10.1093/aje/kwx246

Groves, J. W., Bot, V. A., Ding, D. Y., Nicholas, J., Farinas, A., See-Oh, H., James, S.-N., Wong, A., Williams, D. M., King-Robson, J., Chaturvedi, N., Richards, M., Wyss-Coray, T., & Schott, J. M. (2025). Eight decades of follow-up link life course exposures to proteomic organ ageing and longevity. medRxiv: The Preprint Server for Health Sciences, 2025.09.07.25335188. 10.1101/2025.09.07.25335188

Guo, Y., You, J., Zhang, Y., Liu, W.-S., Huang, Y.-Y., Zhang, Y.-R., Zhang, W., Dong, Q., Feng, J.-F., Cheng, W., & Yu, J.-T. (2024). Plasma proteomic profiles predict future dementia in healthy adults. Nature Aging, 4(2), 247–260. 10.1038/s43587-023-00565-0

Hansson, O., Blennow, K., Zetterberg, H., & Dage, J. (2023). Blood biomarkers for Alzheimer’s disease in clinical practice and trials. Nature Aging, 3(5), 506–519. 10.1038/s43587-023-00403-3

Jack Jr., C. R., Andrews, J. S., Beach, T. G., Buracchio, T., Dunn, B., Graf, A., Hansson, O., Ho, C., Jagust, W., McDade, E., Molinuevo, J. L., Okonkwo, O. C., Pani, L., Rafii, M. S., Scheltens, P., Siemers, E., Snyder, H. M., Sperling, R., Teunissen, C. E., & Carrillo, M. C. (2024). Revised criteria for diagnosis and staging of Alzheimer’s disease: Alzheimer’s Association Workgroup. Alzheimer’s & Dementia, 20(8), 5143–5169. 10.1002/alz.13859

Jackson, R. J., Hyman, B. T., & Serrano-Pozo, A. (2024). Multifaceted roles of APOE in Alzheimer disease. Nature Reviews Neurology, 20(8), 457–474. 10.1038/s41582-024-00988-2

Kirsher, D. Y., Chand, S., Phong, A., Nguyen, B., Szoke, B. G., & Ahadi, S. (2025). Current landscape of plasma proteomics from technical innovations to biological insights and biomarker discovery. Communications Chemistry, 8(1), 279. 10.1038/s42004-025-01665-1

Kuh, D., Wong, A., Shah, I., Moore, A., Popham, M., Curran, P., Davis, D., Sharma, N., Richards, M., Stafford, M., Hardy, R., & Cooper, R. (2016). The MRC National Survey of Health and Development reaches age 70: Maintaining participation at older ages in a birth cohort study. European Journal of Epidemiology, 31(11), 1135–1147. 10.1007/s10654-016-0217-8

Lawler, P. E., Bollinger, J. G., Schindler, S. E., Hodge, C. R., Iglesias, N. J., Krishnan, V., Coulton, J. B., Li, Y., Holtzman, D. M., & Bateman, R. J. (2023). Apolipoprotein E O-glycosylation is associated with amyloid plaques and *APOE* genotype. Analytical Biochemistry, 672, 115156. 10.1016/j.ab.2023.115156

Leckey, C. A., & Zetterberg, H. (2022). Neurofilament light chain: Defining the analyte. Brain Communications, 4(2). 10.1093/braincomms/fcac070

Liu, W.-S., Zhang, Y.-R., Ge, Y.-J., Wang, H.-F., Cheng, W., & Yu, J.-T. (2024). Inflammation and Brain Structure in Alzheimer’s Disease and Other Neurodegenerative Disorders: A Mendelian Randomization Study. Molecular Neurobiology, 61(3), 1593–1604. 10.1007/s12035-023-03648-6

Lumsden, A. L., Mulugeta, A., Zhou, A., & Hyppönen, E. (2020). Apolipoprotein E (APOE) genotype-associated disease risks: A phenome-wide, registry-based, case-control study utilising the UK Biobank. eBioMedicine, 59. 10.1016/j.ebiom.2020.102954

Manichaikul, A., Mychaleckyj, J. C., Rich, S. S., Daly, K., Sale, M., & Chen, W.-M. (2010). Robust relationship inference in genome-wide association studies. Bioinformatics, 26(22), 2867–2873. 10.1093/bioinformatics/btq559

Meng, X., Navoly, G., Giannakopoulou, O., Levey, D. F., Koller, D., Pathak, G. A., Koen, N., Lin, K., Adams, M. J., Rentería, M. E., Feng, Y., Gaziano, J. M., Stein, D. J., Zar, H. J., Campbell, M. L., van Heel, D. A., Trivedi, B., Finer, S., McQuillin, A., … Kuchenbaecker, K. (2024). Multi-ancestry genome-wide association study of major depression aids locus discovery, fine mapping, gene prioritization and causal inference. Nature Genetics, 56(2), 222–233. 10.1038/s41588-023-01596-4

Neuropathology Group. Medical Research Council Cognitive Function and Aging Study. (2001). Pathological correlates of late-onset dementia in a multicentre, community-based population in England and Wales. Neuropathology Group of the Medical Research Council Cognitive Function and Ageing Study (MRC CFAS). Lancet (London, England), 357(9251), 169–175. 10.1016/s0140-6736(00)03589-3

Pereira, J. B., Janelidze, S., Smith, R., Mattsson-Carlgren, N., Palmqvist, S., Teunissen, C. E., Zetterberg, H., Stomrud, E., Ashton, N. J., Blennow, K., & Hansson, O. (2021). Plasma GFAP is an early marker of amyloid-β but not tau pathology in Alzheimer’s disease. Brain, 144(11), 3505–3516. 10.1093/brain/awab223

Polsinelli, A. J., Lane, K. A., Manchella, M. K., Logan, P. E., Gao, S., & Apostolova, L. G. (2023). APOE ε4 is associated with earlier symptom onset in LOAD but later symptom onset in EOAD. Alzheimer’s & Dementia, 19(5), 2212–2217. 10.1002/alz.12955

Privé, F., Aschard, H., Ziyatdinov, A., & Blum, M. G. B. (2018). Efficient analysis of large-scale genome-wide data with two R packages: Bigstatsr and bigsnpr. Bioinformatics (Oxford, England), 34(16), 2781–2787. 10.1093/bioinformatics/bty185

Privé, F., Luu, K., Blum, M. G. B., McGrath, J. J., & Vilhjálmsson, B. J. (2020). Efficient toolkit implementing best practices for principal component analysis of population genetic data. Bioinformatics, 36(16), 4449–4457. 10.1093/bioinformatics/btaa520

Raulin, A.-C., Doss, S. V., Trottier, Z. A., Ikezu, T. C., Bu, G., & Liu, C.-C. (2022). ApoE in Alzheimer’s disease: Pathophysiology and therapeutic strategies. Molecular Neurodegeneration, 17(1), 72. 10.1186/s13024-022-00574-4

Reiman, E. M., Arboleda-Velasquez, J. F., Quiroz, Y. T., Huentelman, M. J., Beach, T. G., Caselli, R. J., Chen, Y., Su, Y., Myers, A. J., Hardy, J., Paul Vonsattel, J., Younkin, S. ., Bennett, D. A., De Jager, P. L., Larson, E. B., Crane, P. K., Keene, C. D., Kamboh, M. I., Kofler, J. K., … Jun, G. R. (2020). Exceptionally low likelihood of Alzheimer’s dementia in APOE2 homozygotes from a 5,000-person neuropathological study. Nature Communications, 11(1), 667. 10.1038/s41467-019-14279-8

Rhinn, H., Tatton, N., McCaughey, S., Kurnellas, M., & Rosenthal, A. (2022). Progranulin as a therapeutic target in neurodegenerative diseases. Trends in Pharmacological Sciences, 43(8), 641–652. 10.1016/j.tips.2021.11.015

Sam Choi / GreedyRelated · GitLab. (2020, July 22). GitLab. https://gitlab.com/choishingwan/GreedyRelated

Schoeler, T., Speed, D., Porcu, E., Pirastu, N., Pingault, J.-B., & Kutalik, Z. (2023). Participation bias in the UK Biobank distorts genetic associations and downstream analyses. Nature Human Behaviour, 7(7), 1216–1227. 10.1038/s41562-023-01579-9

Shireby, G., Morris, T. T., Wong, A., Chaturvedi, N., Ploubidis, G. B., Fitzsimmons, E., Goodman, A., Sanchez-Galvez, A., Davies, N. M., Wright, L., & Bann, D. (2025). Data Resource Profile: Genomic data in multiple British birth cohorts (1946–2001)—linkage with health, social, and environmental data from birth to old age. International Journal of Epidemiology, 54(5), dyaf141. 10.1093/ije/dyaf141

Shvetcov, A., Johnson, E. C. B., Winchester, L. M., Walker, K. A., Wilkins, H. M., Thompson, T. G., Rothstein, J. D., Krish, V., Imam, F. B., Burns, J. M., Swerdlow, R. H., Slawson, C., & Finney, C. A. (2025). APOE ε4 carriers share immune-related proteomic changes across neurodegenerative diseases. Nature Medicine, 31(8), 2590–2601. 10.1038/s41591-025-03835-z

Sun, B. B., Chiou, J., Traylor, M., Benner, C., Hsu, Y.-H., Richardson, T. G., Surendran, P., Mahajan, A., Robins, C., Vasquez-Grinnell, S. G., Hou, L., Kvikstad, E. M., Burren, O. S., Davitte, J., Ferber, K. L., Gillies, C. E., Hedman, Å. K., Hu, S., Lin, T., … Whelan, C. D. (2023). Plasma proteomic associations with genetics and health in the UK Biobank. Nature, 622(7982), 329–338. 10.1038/s41586-023-06592-6

Sun, B. B., Maranville, J. C., Peters, J. E., Stacey, D., Staley, J. R., Blackshaw, J., Burgess, S., Jiang, T., Paige, E., Surendran, P., Oliver-Williams, C., Kamat, M. A., Prins, B. P., Wilcox, S. K., Zimmerman, E. S., Chi, A., Bansal, N., Spain, S. L., Wood, A. M., … Butterworth, A. S. (2018). Genomic atlas of the human plasma proteome. Nature, 558(7708), 73–79. 10.1038/s41586-018-0175-2

Van Hout, C. V., Tachmazidou, I., Backman, J. D., Hoffman, J. D., Liu, D., Pandey, A. K., Gonzaga-Jauregui, C., Khalid, S., Ye, B., Banerjee, N., Li, A. H., O’Dushlaine, C., Marcketta, A., Staples, J., Schurmann, C., Hawes, A., Maxwell, E., Barnard, L., Lopez, A., … Baras, A. (2020). Exome sequencing and characterization of 49,960 individuals in the UK Biobank. Nature, 586(7831), 749–756. 10.1038/s41586-020-2853-0

Verma, A., Huffman, J. E., Rodriguez, A., Conery, M., Liu, M., Ho, Y.-L., Kim, Y., Heise, D. A., Guare, L., Panickan, V. A., Garcon, H., Linares, F., Costa, L., Goethert, I., Tipton, R., Honerlaw, J., Davies, L., Whitbourne, S., Cohen, J., … Liao, K. P. (2024). Diversity and scale: Genetic architecture of 2068 traits in the VA Million Veteran Program. Science, 385(6706), eadj1182. 10.1126/science.adj1182

Viechtbauer, W. (2010). Conducting Meta-Analyses in R with the metafor Package. Journal of Statistical Software, 36, 1–48. 10.18637/jss.v036.i03

Wang, X.-M., Zeng, P., Fang, Y.-Y., Zhang, T., & Tian, Q. (2021). Progranulin in neurodegenerative dementia. Journal of Neurochemistry, 158(2), 119–137. 10.1111/jnc.15378

Wenham, P., Price, W., & Blundell, G. (1991). Apolipoprotein E genotyping by one-stage PCR. The Lancet, 337(8750), 1158–1159. 10.1016/0140-6736(91)92823-K

Wesenhagen, K. E. J., Gobom, J., Bos, I., Vos, S. J. B., Martinez-Lage, P., Popp, J., Tsolaki, M., Vandenberghe, R., Freund-Levi, Y., Verhey, F., Lovestone, S., Streffer, J., Dobricic, V., Bertram, L., Alzheimer’s Disease Neuroimaging Initiative, Blennow, K., Pikkarainen, M., Hallikainen, M., Kuusisto, J., … Tijms, B. M. (2022). Effects of age, amyloid, sex, and APOE ε4 on the CSF proteome in normal cognition. Alzheimer’s & Dementia (Amsterdam, Netherlands), 14(1), e12286. 10.1002/dad2.12286

Williams, D. M., Heikkinen, S., Hiltunen, M., Davies, N. M., & Anderson, E. L. (2026). The proportion of Alzheimer’s disease attributable to apolipoprotein E. Npj Dementia, 2(1), 1. 10.1038/s44400-025-00045-9

Wolters, F. J., Yang, Q., Biggs, M. L., Jakobsdottir, J., Li, S., Evans, D. S., Bis, J. C., Harris, T. B., Vasan, R. S., Zilhao, N. R., Ghanbari, M., Ikram, M. A., Launer, L., Psaty, B. M., Tranah, G. J., Kulminski, A. M., Gudnason, V., Seshadri, S., & E2-CHARGE investigators. (2019). The impact of APOE genotype on survival: Results of 38,537 participants from six population-based cohorts (E2-CHARGE). PloS One, 14(7), e0219668. 10.1371/journal.pone.0219668

Yamazaki, Y., Zhao, N., Caulfield, T. R., Liu, C.-C., & Bu, G. (2019). Apolipoprotein E and Alzheimer disease: Pathobiology and targeting strategies. Nature Reviews Neurology, 15(9), 501–518. 10.1038/s41582-019-0228-7

Yang, L. G., March, Z. M., Stephenson, R. A., & Narayan, P. S. (2023). Apolipoprotein E in lipid metabolism and neurodegenerative disease. Trends in Endocrinology and Metabolism: TEM, 34(8), 430–445. 10.1016/j.tem.2023.05.002

Zetterberg, H., & Schott, J. M. (2022). Blood biomarkers for Alzheimer’s disease and related disorders. Acta Neurologica Scandinavica, 146(1), 51–55. 10.1111/ane.13628

Zhan, H., Cammann, D., Cummings, J. L., Dong, X., & Chen, J. (2025). Biomarker identification for Alzheimer’s disease through integration of comprehensive Mendelian randomization and proteomics data. Journal of Translational Medicine, 23(1), 278. 10.1186/s12967-025-06317-5

Zhang, C., Xie, S., & Malek, M. (2025). SNAP-25: A biomarker of synaptic loss in neurodegeneration. Clinica Chimica Acta, 571, 120236. 10.1016/j.cca.2025.120236

Zhang, L., Xiang, X., Li, Y., Bu, G., & Chen, X.-F. (2025). TREM2 and sTREM2 in Alzheimer’s disease: From mechanisms to therapies. Molecular Neurodegeneration, 20(1), 43. 10.1186/s13024-025-00834-z

