## Supplementary materials: Methods, Results and Figures for "Plasma proteomics of *APOE* genotype: age-specific analyses in UK population-based cohorts"

**Supplementary materials: Plasma proteomic signatures of *APOE* genotype in the UK Biobank**

### Supplementary Method 1: Sample (UK Biobank)

UK Biobank (UKB) is a prospective, multicentre cohort study with deep phenotypic and genetic data collected on approximately 502,000 individuals aged 39-73 years old at baseline, recruited across England, Scotland, and Wales between 2006 and 2010 (Bycroft et al., 2018). Full details of the UKB design, participants, quality control and its strengths and limitations have been described previously (Bycroft et al., 2018; Fry et al., 2017; Schoeler et al., 2023). UKB received initial ethical approval and renewals from the North-West Multi-centre Research Ethics Committee (REC reference numbers 11/NW/0382 and 16/NW/0274).

The UKB Pharma Proteomics Project (UKB-PPP) conducted proteomic profiling on 54,219 plasma samples from UKB participants collected at baseline assessments (Sun et al., 2023). This included a randomly selected subset of 46,595 participants, 6,376 individuals selected by the UKB-PPP consortium members based on diseases of interest, and 1,268 individuals who participated in the COVID-19 repeat-imaging study at multiple visits ($n$ = 1,230 at baseline). Details of UKB-PPP participant selection and thorough comparisons of the random and non-random sub-cohorts have been described previously (Sun et al., 2023).

Here, we used baseline data from UKB-PPP participants who had quality controlled proteomic and whole-exome sequencing (WES) data available (Van Hout et al., 2020), with the latter used to ascertain *APOE* genotypes. Central quality control was performed on the WES data by Regeneron and included sex discordance, contamination, unresolved duplicate sequences and discordance with microarray genotyping data checks (Van Hout et al., 2020). To enable adjustment for genetic principal components generated using genotype data, we then filtered this sample to individuals with quality-controlled genotype data, performing exclusions for failing sample-level genetic quality control (genetic/phenotypic sex mismatches, UKB recommended exclusions based on heterozygosity or missingness, sex chromosome aneuploidy). Kinship estimates provided by UKB were then used to identify related participant pairs (KING $r^2 \geq 0.0442$) (Manichaikul et al., 2010), and the GreedyRelated algorithm was used to remove one individual from each pair, while minimising the number of exclusions required (*Sam Choi / GreedyRelated · GitLab*, 2020). This resulted in analytical samples of 40,092 European, 1,335 African, 971 South Asian, and 244 East Asian ancestry individuals ($N$ = 42,642 across ancestries). European ancestry was defined as participants who self-identified as 'White British' and have similar genetic ancestry based on a principal components analysis of the genotypes (UKB data field ID 22006). The remaining ancestral groups were defined as described previously (Meng et al., 2024).

#### Proteomic measurement, processing and quality control

Full details of the sample handling and central quality control have been described previously (see, Sun et al., 2023). Briefly, 2,941 protein analytes were measured, capturing 2,923 unique proteins, using the antibody-based Olink Explore 3072 proximity extension assay. Each protein is labelled by Olink with an identifier, which typically corresponds to the gene encoding that protein (linked to UniProt IDs), rather than the name of the protein itself. The original UKB-PPP paper reported that one protein (GLIPR1) had >80% of data failing quality control (Sun et al., 2023) – this protein was therefore excluded from our analysis. During quality control by the cohort coordinators, several data points were excluded across proteins due to their identification as likely sample swaps, outliers, or because they were flagged by quality control or assay warnings based on metrics supplied by Olink. Further information on this can be found in the Supplementary Information of Sun et al. (2023).

Protein data were provided as Normalized Protein eXpression (NPX) values. NPX is Olink's arbitrary unit of relative protein quantification on a $\log_2$ scale. Higher NPX values represent higher protein concentrations.

**Supplementary Method 2: Calculation and selection of ancestry-specific genetic principal components for use as covariates**

We calculated genetic principal components (PCs) within each ancestral group analysed to control for population stratification (i.e., genetic confounding). Genetic PCs were calculated using the bigsnpr R package (Privé et al., 2018), following the guidelines outlined by Privé et al. and the accompanying online tutorials (Privé et al., 2020). In summary, we 1) removed all related pairs of individuals (KING kinship coefficient $\geq$ 0.0442; estimates provided by the UK Biobank), 2) performed principal component analysis using the bed_autoSVD on the unrelated participants only, 3) detected principal component outliers and removed them, 4) recalculated the principal components, and 5) projected the PCs onto the entire cohort using the bed_projectSelfPCA function. We amended the number of PCs ($k$) calculated reflecting the amount of population structure present in each ancestry subgroup ($k$ = 10 in EUR and AFR, $k$ = 50 in SAS).

To determine the number of PCs capturing population structure within each ancestral group, we performed visual inspection of scatter plots of the PC loadings and PC scores, as recommended by Privé et al. (Privé et al., 2020). Based on visual inspection of these plots we determined that the first 5 and 8 PCs captured population structure, in individuals of European (EUR) and African (AFR) ancestry, respectively. The remaining PCs appeared to capture other types of structure (e.g., long-range linkage disequilibrium) and noise. However, within individuals of South Asian (SAS) ancestry, evidence of population structure was present across all 50 PCs, though the proportion of variance explained by later PCs is progressively small. To ensure model parsimony, we therefore performed a visual inspection of the scree plot to determine the number of components to retain in analyses of SAS ancestry individuals. In summary, to control for population stratification in our analyses, we therefore included the first 5, 8 and 10 ancestry-specific PCs as covariates for individuals of EUR, AFR, and SAS ancestry, respectively.

### Supplementary Method 3: Validation of *APOE* ε4- and ε2-protein associations in independent cohort SomaLogic data

To validate key *APOE* ε4- and ε2-associated plasma protein findings (FDR-adjusted $p < .05$) identified in the primary UKB analysis of participants of European ancestry, we analysed data from two independent cohorts profiled using proteomic platforms distinct to UKB. Using age-matched groups, a subset of participants from INTERVAL study (Angelantonio et al., 2017) was used to replicate associations identified in the youngest (40–50 years, $n$ = 1,138) and middle (50–60 years, $n$ = 1,062) age groups, while the Medical Research Council National Survey of Health and Development (NSHD; 1946 British birth cohort) (Kuh et al., 2016) was used to replicate associations observed in the oldest age group (60–70 years, $n$ = 2,621). Both replication cohorts were additionally used to assess age-related trends in *APOE*-protein associations identified in the UKB (FDR-adjusted $p < 0.05$).

#### INTERVAL study

INTERVAL is a cohort of approximately 50,000 participants (blood donors aged 18 and over) nested within a randomised trial evaluating the safety of varying blood donation frequency, as previously described (Angelantonio et al., 2017). Between mid-2012 and mid-2014, blood donors aged 18 years and older were recruited at 25 centres of England's National Health Service Blood and Transplant (NHSBT). Participants were generally in good health because blood donation criteria exclude people with a history of major diseases (such as myocardial infarction, stroke, cancer, HIV, and hepatitis B or C) and those who have had recent illness or infection. In this study, we included 1,378 individuals who had relevant covariate, proteomic, and genotype data (for *APOE* coding) available and were aged between 40-50 years ($n$ = 705) or 50-60 years ($n$ = 673).

##### *Genetic data and APOE coding*

Genotyping, quality control, imputation, and ancestry assignment were performed centrally by the INTERVAL study team (Astle et al., 2016) following established protocols. Imputed SNP dosages were centrally converted to hard-call binary PLINK format. From these data, we derived two binary variables representing *APOE* ε2 and ε4 carrier status based on rs7412 and rs429358, using ε3 homozygotes as the reference group and excluding ε2/ε4 heterozygotes.

##### *Proteomic measurement, processing and quality control*

Plasma protein levels were measured using the SomaLogic SomaScan 4K (v3) in INTERVAL, as previously described (Sun et al., 2018). Protein data provided by INTERVAL had been adjusted for age, sex, time between blood draw and processing (binary: ≤1 day vs. >1 day), and the first three ancestry principal components from multidimensional scaling, as residuals from a linear regression of natural log-transformed protein levels (Sun et al., 2018).

#### NSHD study

The MRC National Survey of Health and Development (NSHD) is a population-based birth cohort of 5,362 individuals born in a single week in March 1946 (Kuh et al., 2016). Of the

5,362 original birth cohort participants, 3,163 were alive and enrolled at a mean age of 63.2 years, when blood samples for plasma proteomics were collected. A total of 1,803 participants attended one of five UK assessment centres (London, Manchester, Birmingham, Cardiff, and Edinburgh) and underwent peripheral venepuncture. In this study, we included 1,645 individuals who had relevant covariate, proteomic, and genotype data (for *APOE* coding) available.

***Genetic data and APOE coding***

Genotyping, quality control, imputation, and ancestry assignment were performed centrally by the NSHD study team (Shireby et al., 2025) following established protocols. Imputed SNP dosages were centrally converted to hard-call binary PLINK format. To determine the number of genetic principal components (PCs) to adjust for in analyses, we sequentially plotted PCs until subsequent components showed no observable association with the outcome. From these data, we derived two binary variables representing *APOE* ε2 and ε4 carrier status based on rs7412 and rs429358, using ε3 homozygotes as the reference group and excluding ε2/ε4 heterozygotes.

***Proteomic measurement, processing and quality control***

Plasma protein levels were measured using the SomaLogic SomaScan 11k (v5.0) (Candia et al., 2024) in NSHD. Although the SomaScan 11K platform includes aptamers for specific *APOE* isoforms (ApoE2, ApoE3, and ApoE4), we excluded them from analyses given evidence of their poor selectivity (Kirsher et al., 2025); total ApoE (non-isoform specific) was retained. Values below the estimated LOD were retained in line with SomaLogic recommendations.

**Statistical analysis**

Overlap between *APOE* ε4- and ε2-associated plasma proteins identified in UKB and those available in the independent cohorts is reported in the Results. Analyses were conducted in the same way as the main UKB analysis where possible. INTERVAL protein data were pre-adjusted residuals accounting for age, sex, sample processing time, and genetic ancestry (Sun et al., 2018); therefore, we did not include additional covariates in our analyses. To assess the impact of this difference in methods, we repeated the UKB analyses after first residualising protein levels for covariates prior to rank-based inverse normal transformation and regression on *APOE* carrier status, observing minimal effects on the results. In NSHD analyses, the same covariates as the main UKB analysis, except in the UKB-PPP sub-cohort, where adjustments were made for age (within age groups), sex, proteomic batch (plate run date), and the first seven genetic PCs. Where multiple SomaScan aptamers mapped to a single UniProt identifier, all were analysed with Bonferroni correction applied ($p$ divided by the number of aptamers per UniProt).

**Ethical approval**

All participants provided written informed consent; ethical approval was granted to INTERVAL by the National Research Ethics Service (11/EE/0538) and to NSHD by the National Research Ethics Service Committee London (14/LO/1173) and Scotland A Research Ethics Committee (14/SS/1009).

**Supplementary Method 4: Sensitivity analyses**

To investigate the robustness of our findings, we conducted the following sensitivity analyses in the primary analytical sample (UKB participants of European ancestry) across all 2,922 available proteins. First, we repeated all analyses using z-scored protein levels within each age group without the rank-based inverse normal transformation applied. We examined the percentage of associations that remained showing evidence of an association without the transformation applied to data (i.e., FDR-adjusted $P$ <.05). We also calculated the Pearson's correlation coefficient ($r$) between effect estimates derived from the transformed and untransformed data across proteins, both in the whole set of results and stratified by carriership comparison (ε4 or ε2 vs. ε3 homozygotes) and by age group. To examine potential differences in *APOE* effects across UKB-PPP selection sub-cohorts, we repeated analyses separately in randomly selected individuals ($n$ = 34,551) and in those selected by the UKB-PPP consortium or the COVID-19 imaging study ($n$ = 5,541). We assessed the percentage of associations that showed evidence of an association in the non-random sub-cohorts that were replicated in the random sub-cohort (i.e., FDR-adjusted $P$ <.05). We also computed Pearson's $r$ between effect estimates from the random and non-random sub-cohorts, across all proteins and for those showing evidence of associations in the random sample (FDR-adjusted), stratifying by carriership comparison (ε4 and ε2 vs. ε3 homozygotes) and age group.

### Supplementary Result 1: Replication of APOE ε4- and ε2-protein associations in independent cohorts using SomaLogic data

To replicate *APOE* ε4- and ε2-associated plasma protein findings identified in UKB (FDR-adjusted $P < 0.05$), we performed approximately age-matched analyses in two independent cohorts, INTERVAL and NSHD, that used SomaLogic plasma proteomic assays (SomaScan 4K v3.0 and 11K v5.0, respectively). Replication was defined as evidence of association ($P < 0.05$) with effects directionally concordant with those observed in UKB. Participant characteristics split by age group and ancestry are reported in Supplementary Table 12. Sample sizes per analysis, including the number of *APOE* ε4 or ε2 carriers in each protein level comparison with ε3 homozygotes, are provided alongside the full results in Supplementary Tables 13-14.

First, in the youngest age group (40-50 years), 59.4% of *APOE*-associated proteins identified in UKB were available in INTERVAL. Four ε4-associated (LDLR, MENT, NAAA, PLA2G7) and one ε2-associated protein (PLA2G7) showed evidence of replication in the INTERVAL cohort. Second, in the middle age group (50-60 years), 55.6% of *APOE*-associated proteins identified in UKB were available in INTERVAL. Six ε4-associated (BOC, CCL25, MENT, MMP8, RTN4R, TREML2) and 10 ε2-associated (CCL23, CCL25, CCN1, CXCL13, GRN, ITGAL, MENT, MMP8, PLA2G7, RTN4R) proteins were replicated in INTERVAL. Third, in the oldest age group (60-70 years in UKB and 60-65 years in NSHD), 92.9% of *APOE*-associated proteins identified in UKB were available for analysis in the NSHD cohort. In total, 25 ε4-associated and 26 ε2-associated proteins were replicated in NSHD. Notably, replicated ε4-associated proteins included seven robustly associated ε4 carrier status in UKB (AGR3, ANGPTL3, APOE, CES1, MENT, NAAA, PLA2G7). Further, replicated ε2-associated proteins included two encoded by known genetic risk loci for AD (CTSB, GRN), seven robustly associated with ε2 carrier status in UKB (APOC1, BRK1, HMOX1, IFGR2, MENT, MXRA8, PLA2G7), and one shown to be robustly associated with ε4 (but not ε2) carrier status in UKB (AGR3). Associations for NEFL in ε4 carriers were directionally opposite to those observed in UKB, consistent with prior reports of cross-platform discordance and inverse associations with AD risk when measured using SomaScan (Budelier et al., 2022; Frick et al., 2024; Leckey & Zetterberg, 2022). Data on NEFL levels were not available in the INTERVAL study. Similarly, SNAP25 showed strong positive associations in ε4 carriers within UKB, whereas inverse associations were observed across replication cohorts, potentially reflecting platform-specific measurement differences. Related, ε4 carriers in the oldest UKB age group exhibited lower GRN levels; however, a positive direction of effect was observed in NSHD. Full NSHD replication results are provided in Supplementary Tables 13-14 for ε4- and ε2-associated proteins, respectively.

For age-related trends, 14 of the 25 proteins identified in UKB (56.0%) among ε4 carriers were available across age groups in INTERVAL and NSHD. Directional concordance with UKB was observed for 7 of 15 corresponding aptamers, although all *P* values for trends in the independent cohorts exceeded 0.05 (Supplementary Table 15). No age-associated protein trends were detected among *APOE* ε2 carriers in UKB, and therefore, no replication analyses were conducted for this group.

### Supplementary Result 2: Sensitivity analyses

Most protein levels showing evidence of an association with *APOE* ε4 or ε2 (*P* <.05, FDR-adjusted) using rank-based inverse normal transformation also showed evidence of an association with the same direction of effect in analyses without the transformation applied (91.2% for ε4, 81.7% for ε2, across age groups). Effect estimates were highly correlated between the two approaches (Supplementary Figure 3; full results in Supplementary Tables 16–17; correlations by age group in Supplementary Table 18). Most proteins showing evidence of an association with *APOE* ε4 or ε2 (*p* <.05, FDR-adjusted) in the smaller, non-randomly selected UKB-PPP sample also showed evidence of an association in the same direction in randomly selected participants (64.1% for ε4, 95.0% for ε2). Effect estimates across all proteins were weakly to moderately correlated between random and non-random participants, except for ε4 carriers in the oldest age group, where a strong correlation was observed. Restricting to proteins associated with *APOE* ε4 or ε2 in at least one age group in the non-random sample yielded very strong correlations for ε4, and strong to very strong correlations for ε2 carriers (Supplementary Figure 4; full results in Supplementary Tables 19–22; correlations by age group in Supplementary Table 23).

**Supplementary Figure 1. Estimated effects for proteins robustly associated with *APOE* genotype across multiple age groups in African ancestry individuals.**

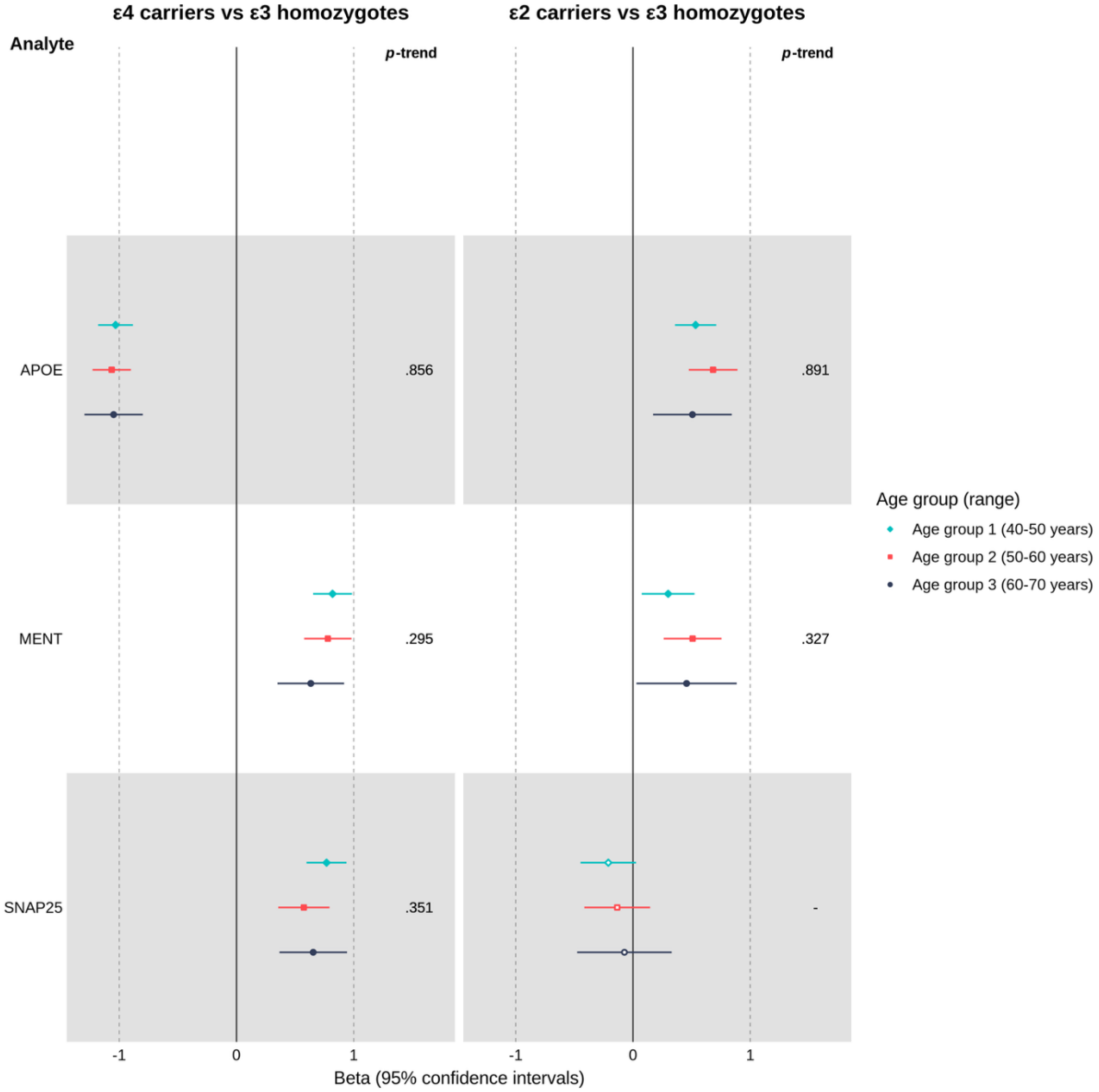

***Note.*** This forest plot shows the effect estimates (betas) and 95% confidence intervals for proteins that were robustly associated with *APOE* genotype ($p < .001$, unadjusted) in two or more age groups in comparisons of either *APOE* ε4 or ε2 carriers with ε3 homozygotes of African ancestry. All proteins (APOE, MENT, SNAP25) shown met these criteria among ε4 carriers, and APOE met these criteria among ε2 carriers. Betas represent the standardised mean difference in inverse-normal transformed protein levels (in SD units) between carriers and non-carriers, adjusting for covariates. Filled points represent beta estimates with $p < .05$ (unadjusted). The left panel shows results for ε4 vs. ε3 homozygotes, whereas the right panel shows the results for the same proteins in ε2 vs. ε3 homozygotes. On the right-hand side of each panel is the *p*-value indicating whether there was evidence of an age-trend in associations (all *ps* > .05, unadjusted) for proteins showing evidence of an association in at least one age group.

**Supplementary Figure 2. Estimated effects for proteins robustly associated with APOE ε4 carrier status across multiple age groups in South Asian ancestry individuals.**

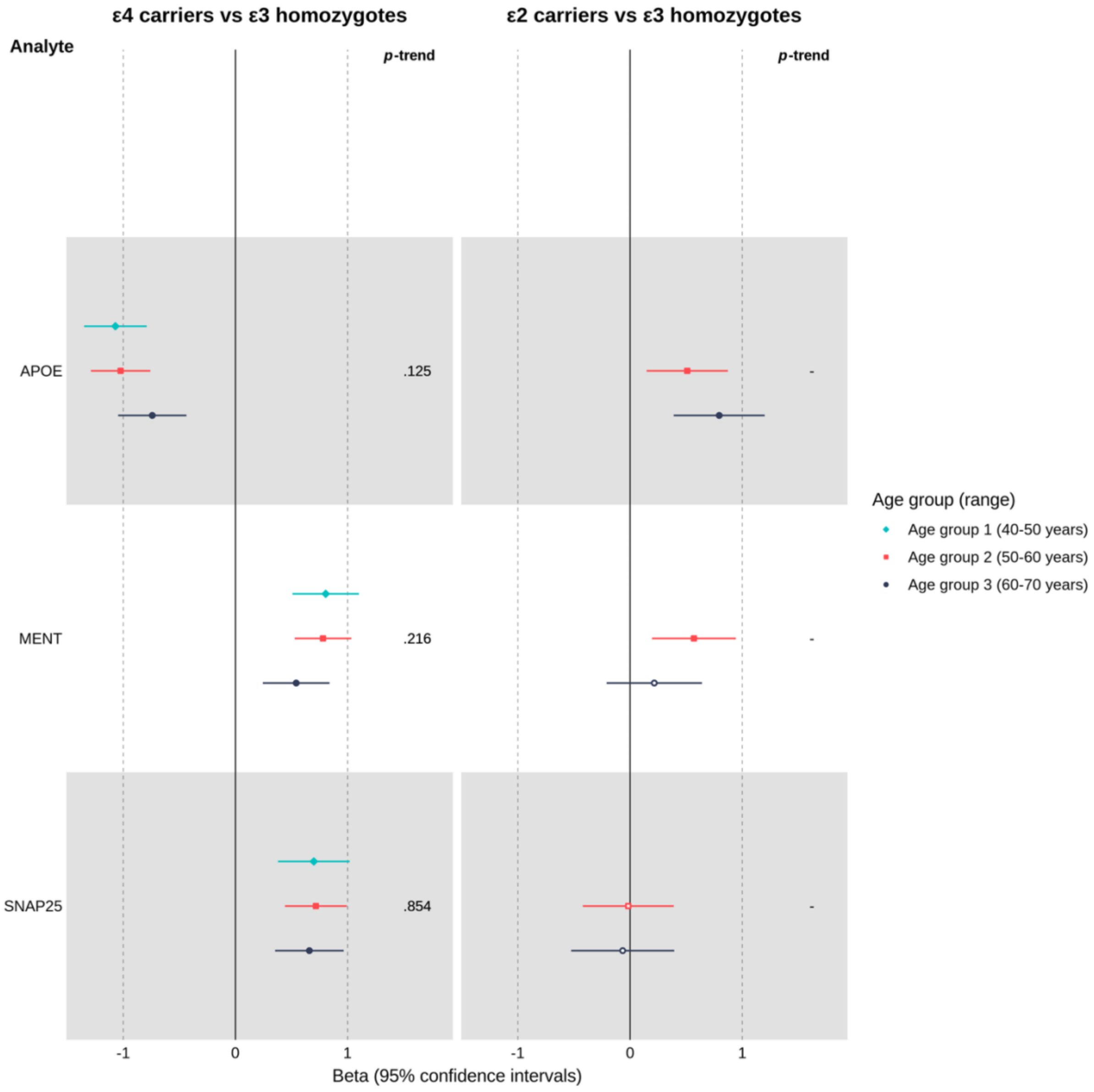

***Note.*** This forest plot displays effect estimates (betas) and 95% confidence intervals for proteins robustly associated with APOE genotype ($p < 0.001$, unadjusted) in two or more age groups among individuals of South Asian ancestry. Proteins meeting these criteria were observed for ε4 versus ε3 homozygotes (APOE, MENT, SNAP25), whereas no proteins met the criteria for ε2 versus ε3 homozygotes. Betas represent the standardised mean difference in inverse-normal–transformed protein levels (SD units) between carriers and non-carriers, adjusted for covariates; filled points indicate $p < 0.05$ (unadjusted). The left panel shows ε4 vs. ε3 results, and the right panel shows corresponding ε2 vs. ε3 results. Results for ε2 vs. ε3 homozygotes are not shown in the youngest age group due to insufficient sample size. P-values on the right of each panel indicate evidence for age-related trends (all *ps* > 0.05, unadjusted).

**Supplementary Figure 3. Scatter plot of effect sizes from protein data with and without inverse normal transformation**

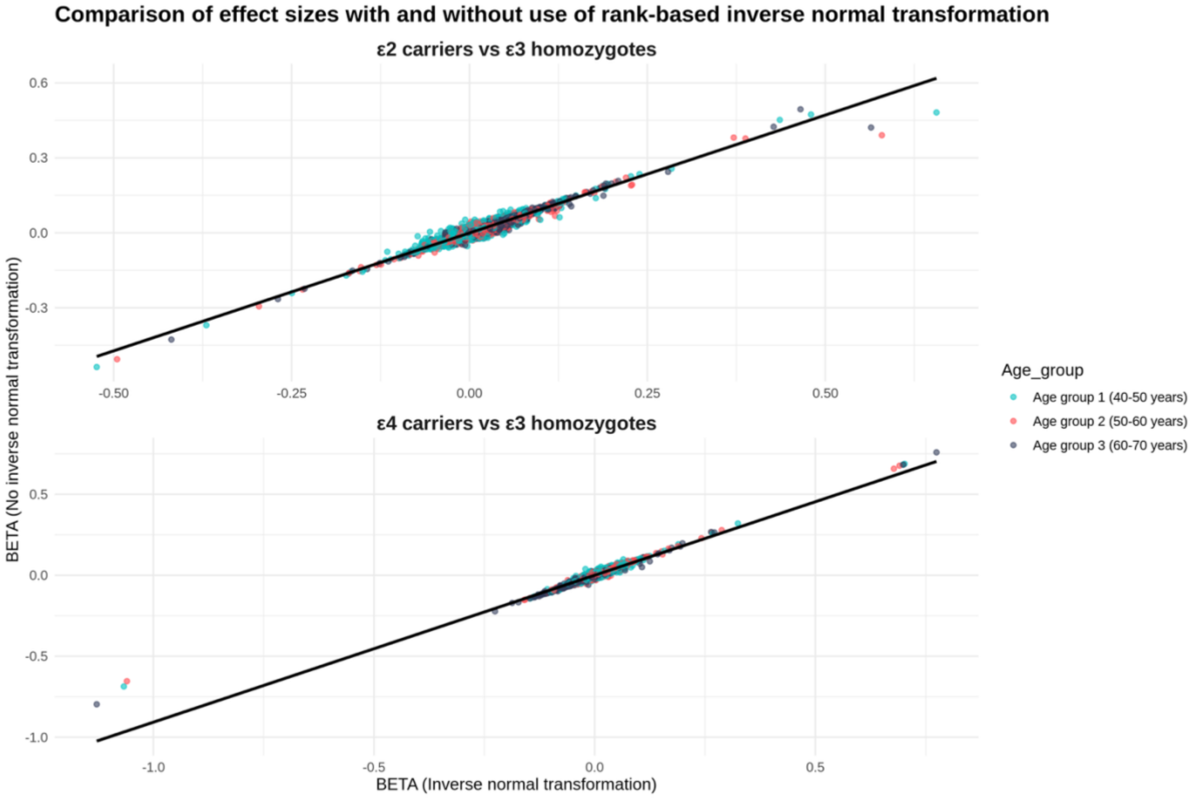

***Note.*** This scatter plot shows the effects estimates (betas) from analyses with and without rank-based inverse normal transformation applied to protein level outcome data in participants of European ancestry (*N* = 40,092; sample sizes per model vary by ε2/ε4 carriership comparison and protein due to quality control procedures and missing data). The top panel shows the effect estimates for *APOE* ε2 carriers vs. ε3 homozygotes, whereas the bottom panel shows *APOE* ε4 carriers vs. ε3 homozygotes.

**Supplementary Figure 4. Scatter plot of effect sizes from randomly selected participants vs. those selected by the UKB-PPP consortium or the COVID-19 imaging study**

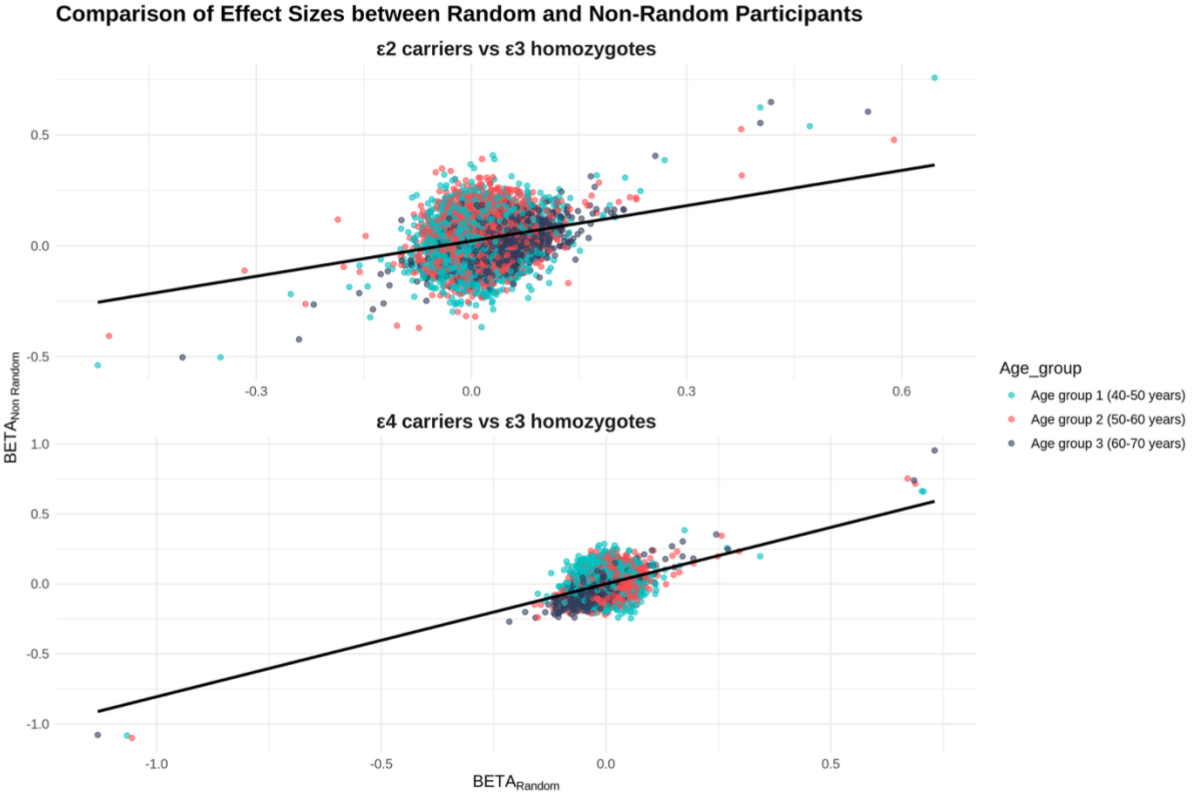

***Note.*** UKB-PPP = UK Biobank Pharma Proteomics Project. This scatter plot shows the effects estimates (betas) from analyses in randomly selected participants European ancestry (*n* = 34,551 and *n* = 5,541, respectively; sample sizes per model vary by ε2/ε4 carriership comparison and protein due to quality control procedures and missing data). The top panel shows the effect estimates for *APOE* ε2 carriers vs. ε3 homozygotes, whereas the bottom panel shows *APOE* ε4 carriers vs. ε3 homozygotes.

### Supplementary References

Angelantonio, E. D., Thompson, S. G., Kaptoge, S., Moore, C., Walker, M., Armitage, J., Ouwehand, W. H., Roberts, D. J., Danesh, J., Angelantonio, E. D., Thompson, S. G., Kaptoge, S., Moore, C., Walker, M., Armitage, J., Ouwehand, W. H., Roberts, D. J., Danesh, J., Armitage, J., … Roberts, D. J. (2017). Efficiency and safety of varying the frequency of whole blood donation (INTERVAL): A randomised trial of 45 000 donors. *The Lancet*, *390*(10110), 2360–2371. https://doi.org/10.1016/S0140-6736(17)31928-1

Astle, W. J., Elding, H., Jiang, T., Allen, D., Ruklisa, D., Mann, A. L., Mead, D., Bouman, H., Riveros-Mckay, F., Kostadima, M. A., Lambourne, J. J., Sivapalaratnam, S., Downes, K., Kundu, K., Bomba, L., Berentsen, K., Bradley, J. R., Daugherty, L. C., Delaneau, O., … Soranzo, N. (2016). The Allelic Landscape of Human Blood Cell Trait Variation and Links to Common Complex Disease. *Cell*, *167*(5), 1415-1429.e19. https://doi.org/10.1016/j.cell.2016.10.042

Budelier, M. M., He, Y., Barthelemy, N. R., Jiang, H., Li, Y., Park, E., Henson, R. L., Schindler, S. E., Holtzman, D. M., & Bateman, R. J. (2022). A map of neurofilament light chain species in brain and cerebrospinal fluid and alterations in Alzheimer’s disease. *Brain Communications*, *4*(2). https://doi.org/10.1093/braincomms/fcac045

Bycroft, C., Freeman, C., Petkova, D., Band, G., Elliott, L. T., Sharp, K., Motyer, A., Vukcevic, D., Delaneau, O., O’Connell, J., Cortes, A., Welsh, S., Young, A., Effingham, M., McVean, G., Leslie, S., Allen, N., Donnelly, P., & Marchini, J. (2018). The UK Biobank resource with deep phenotyping and genomic data. *Nature*, *562*(7726), 203–209. https://doi.org/10.1038/s41586-018-0579-z

Candia, J., Fantoni, G., Delgado-Peraza, F., Shehadeh, N., Tanaka, T., Moaddel, R., Walker, K. A., & Ferrucci, L. (2024). Variability of 7K and 11K SomaScan Plasma Proteomics

Assays. *Journal of Proteome Research*, *23*(12), 5531–5539. https://doi.org/10.1021/acs.jproteome.4c00667

Frick, E. A., Emilsson, V., Jonmundsson, T., Steindorsdottir, A. E., Johnson, E. C. B., Puerta, R., Dammer, E. B., Shantaraman, A., Cano, A., Boada, M., Valero, S., García-González, P., Gudmundsson, E. F., Gudjonsson, A., Pitts, R., Qiu, X., Finkel, N., Loureiro, J. J., Orth, A. P., … Gudnason, V. (2024). Serum proteomics reveal APOE-ε4-dependent and APOE-ε4-independent protein signatures in Alzheimer’s disease. *Nature Aging*, *4*(10), 1446–1464. https://doi.org/10.1038/s43587-024-00693-1

Fry, A., Littlejohns, T. J., Sudlow, C., Doherty, N., Adamska, L., Sprosen, T., Collins, R., & Allen, N. E. (2017). Comparison of Sociodemographic and Health-Related Characteristics of UK Biobank Participants With Those of the General Population. *American Journal of Epidemiology*, *186*(9), 1026–1034. https://doi.org/10.1093/aje/kwx246

Kirsher, D. Y., Chand, S., Phong, A., Nguyen, B., Szoke, B. G., & Ahadi, S. (2025). Current landscape of plasma proteomics from technical innovations to biological insights and biomarker discovery. *Communications Chemistry*, *8*(1), 279. https://doi.org/10.1038/s42004-025-01665-1

Kuh, D., Wong, A., Shah, I., Moore, A., Popham, M., Curran, P., Davis, D., Sharma, N., Richards, M., Stafford, M., Hardy, R., & Cooper, R. (2016). The MRC National Survey of Health and Development reaches age 70: Maintaining participation at older ages in a birth cohort study. *European Journal of Epidemiology*, *31*(11), 1135–1147. https://doi.org/10.1007/s10654-016-0217-8

Leckey, C. A., & Zetterberg, H. (2022). Neurofilament light chain: Defining the analyte. *Brain Communications*, *4*(2). https://doi.org/10.1093/braincomms/fcac070

Manichaikul, A., Mychaleckyj, J. C., Rich, S. S., Daly, K., Sale, M., & Chen, W.-M. (2010). Robust relationship inference in genome-wide association studies. *Bioinformatics*, *26*(22), 2867–2873. https://doi.org/10.1093/bioinformatics/btq559

Meng, X., Navoly, G., Giannakopoulou, O., Levey, D. F., Koller, D., Pathak, G. A., Koen, N., Lin, K., Adams, M. J., Rentería, M. E., Feng, Y., Gaziano, J. M., Stein, D. J., Zar, H. J., Campbell, M. L., van Heel, D. A., Trivedi, B., Finer, S., McQuillin, A., … Kuchenbaecker, K. (2024). Multi-ancestry genome-wide association study of major depression aids locus discovery, fine mapping, gene prioritization and causal inference. *Nature Genetics*, *56*(2), 222–233. https://doi.org/10.1038/s41588-023-01596-4

Privé, F., Aschard, H., Ziyatdinov, A., & Blum, M. G. B. (2018). Efficient analysis of large-scale genome-wide data with two R packages: Bigstatsr and bigsnpr. *Bioinformatics (Oxford, England)*, *34*(16), 2781–2787. https://doi.org/10.1093/bioinformatics/bty185

Privé, F., Luu, K., Blum, M. G. B., McGrath, J. J., & Vilhjálmsson, B. J. (2020). Efficient toolkit implementing best practices for principal component analysis of population genetic data. *Bioinformatics*, *36*(16), 4449–4457. https://doi.org/10.1093/bioinformatics/btaa520

*Sam Choi / GreedyRelated · GitLab*. (2020, July 22). GitLab. https://gitlab.com/choishingwan/GreedyRelated

Schoeler, T., Speed, D., Porcu, E., Pirastu, N., Pingault, J.-B., & Kutalik, Z. (2023). Participation bias in the UK Biobank distorts genetic associations and downstream analyses. *Nature Human Behaviour*, *7*(7), 1216–1227. https://doi.org/10.1038/s41562-023-01579-9

Shireby, G., Morris, T. T., Wong, A., Chaturvedi, N., Ploubidis, G. B., Fitzsimmons, E., Goodman, A., Sanchez-Galvez, A., Davies, N. M., Wright, L., & Bann, D. (2025). Data Resource Profile: Genomic data in multiple British birth cohorts (1946–2001)—linkage with health, social, and environmental data from birth to old age. *International Journal of Epidemiology*, *54*(5), dyaf141. https://doi.org/10.1093/ije/dyaf141

Sun, B. B., Chiou, J., Traylor, M., Benner, C., Hsu, Y.-H., Richardson, T. G., Surendran, P., Mahajan, A., Robins, C., Vasquez-Grinnell, S. G., Hou, L., Kvikstad, E. M., Burren, O. S., Davitte, J., Ferber, K. L., Gillies, C. E., Hedman, Å. K., Hu, S., Lin, T., … Whelan, C. D. (2023). Plasma proteomic associations with genetics and health in the UK Biobank. *Nature*, *622*(7982), 329–338. https://doi.org/10.1038/s41586-023-06592-6

Sun, B. B., Maranville, J. C., Peters, J. E., Stacey, D., Staley, J. R., Blackshaw, J., Burgess, S., Jiang, T., Paige, E., Surendran, P., Oliver-Williams, C., Kamat, M. A., Prins, B. P., Wilcox, S. K., Zimmerman, E. S., Chi, A., Bansal, N., Spain, S. L., Wood, A. M., … Butterworth, A. S. (2018). Genomic atlas of the human plasma proteome. *Nature*, *558*(7708), 73–79. https://doi.org/10.1038/s41586-018-0175-2

Van Hout, C. V., Tachmazidou, I., Backman, J. D., Hoffman, J. D., Liu, D., Pandey, A. K., Gonzaga-Jauregui, C., Khalid, S., Ye, B., Banerjee, N., Li, A. H., O’Dushlaine, C., Marcketta, A., Staples, J., Schurmann, C., Hawes, A., Maxwell, E., Barnard, L., Lopez, A., … Baras, A. (2020). Exome sequencing and characterization of 49,960 individuals in the UK Biobank. *Nature*, *586*(7831), 749–756. https://doi.org/10.1038/s41586-020-2853-0
